# The metabolic, virulence and antimicrobial resistance profiles of colonizing *Streptococcus pneumoniae* shift after pneumococcal vaccine introduction in urban Malawi

**DOI:** 10.1101/2021.07.21.21260914

**Authors:** Andrea Gori, Uri Obolski, Todd D. Swarthout, José Lourenço, Caroline M. Weight, Jen Cornick, Arox Kamng’ona, Thandie S. Mwalukomo, Jacquline Msefula, Comfort Brown, Martin C. Maiden, Neil French, Sunetra Gupta, Robert S. Heyderman

## Abstract

*Streptococcus pneumoniae* accounts for at least 300,000 deaths from pneumonia, septicaemia and meningitis among children under 5-years-old worldwide. Protein–polysaccharide conjugate vaccines (PCVs) are highly effective at reducing vaccine serotype disease but emergence of non-vaccine serotypes and persistent nasopharyngeal carriage threaten to undermine this success. Here, we address the hypothesis that following vaccine introduction in high disease and carriage burden settings, adapted pneumococcal genotypes emerge with the potential to facilitate vaccine escape. We show that beyond serotype replacement, there are marked changes in *S. pneumoniae* carriage population genetics amongst 2804 isolates sampled 4-8 years after the 2011 introduction of PCV-13 in urban Malawi. These changes are characterised by metabolic genotypes with distinct virulence and antimicrobial resistance (AMR) profiles. This included exclusive genes responsible for metabolism and carbohydrate transport, and toxin-antitoxin systems located in an integrative-conjugative region suggestive of horizontal gene transfer. These emergent genotypes were found to have differential growth, haemolytic, or epithelial adhesion/invasion traits that may confer advantage in the nasopharyngeal niche. Together these data show that in the context of PCV13 introduction in a high burden population, there has been a shift in the pneumococcal population dynamics with the emergence of genotypes that have undergone multiple adaptations extending beyond simple serotype replacement, a process that could further undermine vaccine control and promote the spread of AMR.

## Introduction

*Streptococcus pneumoniae* (the pneumococcus), a common commensal of the upper respiratory tract, is responsible for a high burden of severe pneumonia, septicaemia and meningitis [O’Brien, *et al*., 2013], mainly affecting children under 5-years-old, the immunocompromised and the elderly. Pneumococcal disease causes almost 300,000 deaths annually in under-5’s worldwide, and disproportionally affects people in resource-poor settings, where 57% of the total pneumococcal deaths occur [Wahl *et al*., 2015]. This high burden of pneumococcal disease is largely vaccine preventable [O’ Brien, *et al*. 2009; O’ Brien 2013; Ngocho, *et al*., 2019]. Indeed, reports from The Gambia, South Africa, Kenya and Malawi [Roca, *et al*., 2015; Nzenze, S. A. *et al*. 2015; Cohen *et al*., 2017; Hammitt *et al*., 2019, Bar-Zeev, *et al*., 2021] show that pneumococcal conjugate vaccines (PCV), targeting the polysaccharide capsule of the most common disease-causing pneumococcal serotypes (currently PCV10 or PCV13), have been highly effective in reducing invasive pneumococcal disease (IPD). However, the highly adaptable nature of *S. pneumoniae* has the potential to undermine this success. Through the expression of approximately 100 capsule serotypes, the pneumococcus has the ability to adapt to change, by mutation and by the capacity to readily acquire new traits through horizontal gene transfer from other pneumococci and related streptococci occupying the same niche [Salvadori *et al*., 2019].

In the context of PCV introduction, vaccine type (VT) replacement IPD is widespread in resource-rich settings [Southern, *et al*., 2018; Vestrheim, *et al*., 2010] and is being increasingly reported on the African continent [Kwambana-Adams, *et al*., 2017, Vadlamudi, *et al*., 2019]. In addition, we and others have shown that despite excellent vaccine uptake in many African countries, there is considerable residual VT nasopharyngeal carriage in both PCV-vaccinated and PCV-unvaccinated populations several years after PCV introduction [Swarthout, *et al*., 2020, Roca, *et al*., 2015; Nzenze, S. A. *et al*. 2015; Cohen, *et al*., 2017]. Routine PCV programmes have also been shown to reduce the burden of antimicrobial resistant (AMR) *S. pneumoniae* VT disease and carriage in some settings [Ben-Shimol, *et al*., 2018; Tin Htar, *et al*., 2019]. However, the impact has not been uniform, with the emergence of certain AMR serotypes, most notably 7F and 19A [Lee, *et al*., 2017; Ghahfarokhi, *et al*., 2020; Lo, *et al*., 2019]. Furthermore, our modelling suggests that under vaccine pressure, antimicrobial resistant non-VT (NVT) strains could replace susceptible NVT strains through the removal of competition from vaccine-susceptible VTs [Obolski *et al*., 2018].

Emerging evidence from Africa, Europe and the US suggests that PCV introduction has driven genomic alterations in *S. pneumoniae* populations, contributing to capsular switching, selection of specific lineages and genetic recombination [Chaguza, *et al*., 2018; Azarian *et al*., 2018; Croucher, *et al*., 2014; Croucher, *et al*., 2015; Sheppard, *et al*., 2019]. We have previously described a theoretical framework whereby pneumococcal vaccines targeting particular serotypes drive the emergence of NVT *S. pneumoniae* strains with metabolic and virulence-associated characteristics similar to the VTs commonly circulating prior to vaccine introduction [Watkins, *et al*., 2015].

Here, to explore the genotypic and phenotypic basis for this framework, we have used a large, well-characterised pneumococcal carriage strain collection from an urban population in Blantyre, Malawi (2015-2018), established starting 4 years after the introduction PCV13 into the national immunisation programme. PCV13 was introduced 12 November 2011 using a 3+0 schedule, with primary doses given at 6, 10 and 14 weeks of age. Field studies in Malawi have reported high PCV13 uptake of 90%– 95% [Mvula, *et al.,* 2016], similar to the 92% PCV13 coverage recently reported by WHO/UNICEF. [WHO and UNICEF estimates of immunization coverage, 2018. https://www.who.int/immunization/monitoring_surveillance/routine/coverage/WUENIC_notes.pdf] We hypothesised that in this population with a high force of infection [Lourenço, *et al*. 2019] and a high frequency of multiple serotype carriage [Swarthout, *et al.,* 2020], there would be an increase in pneumococcal diversity, adaptability and potentially AMR. We have therefore developed a metabolic core genome allelic profiling method, which has allowed us to analyse 2804 *S. pneumoniae* carriage isolates collected over 4 years of surveillance. We show that discreet “metabolic genotypes” have expanded amongst both VT and NVT serotypes up to 7 years after vaccine introduction. These expanding metabolic genotypes are characterised by accessory genes linked to virulence and AMR, as well as *in-vitro* phenotypes that suggest adaptation. Together these data highlight mechanisms of vaccine escape beyond serotype replacement that, if not addressed, may undermine current and future vaccine strategies.

## Results

### Homogeneity of the Blantyre pneumococcal carriage strain collection across age-cohorts using classical serotyping and genotyping methods

In our population-based carriage surveillance in Blantyre, Malawi (8 surveys between 2015-2019), we have shown high persistent residual VT carriage among PCV-vaccinated children 3–5-year-old (16.7%) and PCV-unvaccinated children 6–8-year-old (15.7%) and HIV-infected adults 18-40 years old on antiretroviral therapy (ART; 8.9%) [Swarthout, *et al*., 2020]. Using this Blantyre pneumococcal carriage strain collection, WGS libraries were developed using a random subset of isolates from each of the eight surveys (Table 1).

**Table 1.** Number of isolates, sequenced per community carriage survey. Surveys were conducted every 6 months, commencing in June 2015. Columns show the number of sequenced isolates and the percent of carriage identified in each of the 3 cohorts included in this study.

| Survey # | Survey dates | Vaccinated children (2-7 y/o) |  | Unvaccinated children (5-10 y/o) |  | Adults (18-40 y/o HIV+ on ART) |  |
| --- | --- | --- | --- | --- | --- | --- | --- |
|  |  | Sequenced isolates # | % carriage | Sequenced isolates # | % carriage | Sequenced isolates # | % carriage |
| **1** | June 2015<br>August 2015 | 222 | 84.3 | 133 | 67.9 | 71 | 39.4 |
| **2** | October 2015<br>April 2016 | 200 | 75.9 | 128 | 62.3 | 69 | 47.2 |
| **3** | May 2016<br>October 2016 | 245 | 78.1 | 114 | 58.7 | N/A | 44.5 |
| **4** | November 2016<br>April 2017 | 226 | 69.6 | 71 | 37.9 | N/A | 42.9 |
| **5** | May 2017<br>October 2017 | 215 | 81 | 45 | 58.5 | 64 | 38.4 |
| **6** | November 2017<br>June 2018 | 219 | 63.9 | 62 | 48.5 | 51 | 32.5 |
| **7** | June 2018<br>December 2018 | 253 | 77.4 | 80 | 57.4 | N/A | 38.6 |
| **8** | January 2019<br>June 2019 | 269 | 68.2 | 67 | 48.2 | N/A | 32.9 |

Among the final dataset of 2804 sequenced carriage isolates, we observed a decline of vaccine-type serotype 1 and 6B carriage isolates, and an increase in frequency amongst several NVTs over time, in particular 23B and 38 (Figure 1). However, the majority of VTs and NVTs persisted over-time and were equally distributed across age cohorts (Figure 1, Figure S1). Two hundred sequence-types (ST) were identified on the basis of the allelic profiles of a standard set of seven housekeeping genes (see Methods). Most STs were present in each cohort (Figure S1a), with the exception of the STs circulating at very low frequencies, including ST347, ST4084, ST10880 and ST10992, which were not identified among the isolates from the HIV-infected adults. Similarly, identified serotypes and global pneumococcal sequencing clusters (GPSCs) were evenly distributed across the age-cohorts surveyed (figure S1b and S1c). Given this spread of serotyping and genotyping characteristics throughout the collection, all isolates were aggregated into a single dataset for the subsequent analyses.

**Figure 1.**
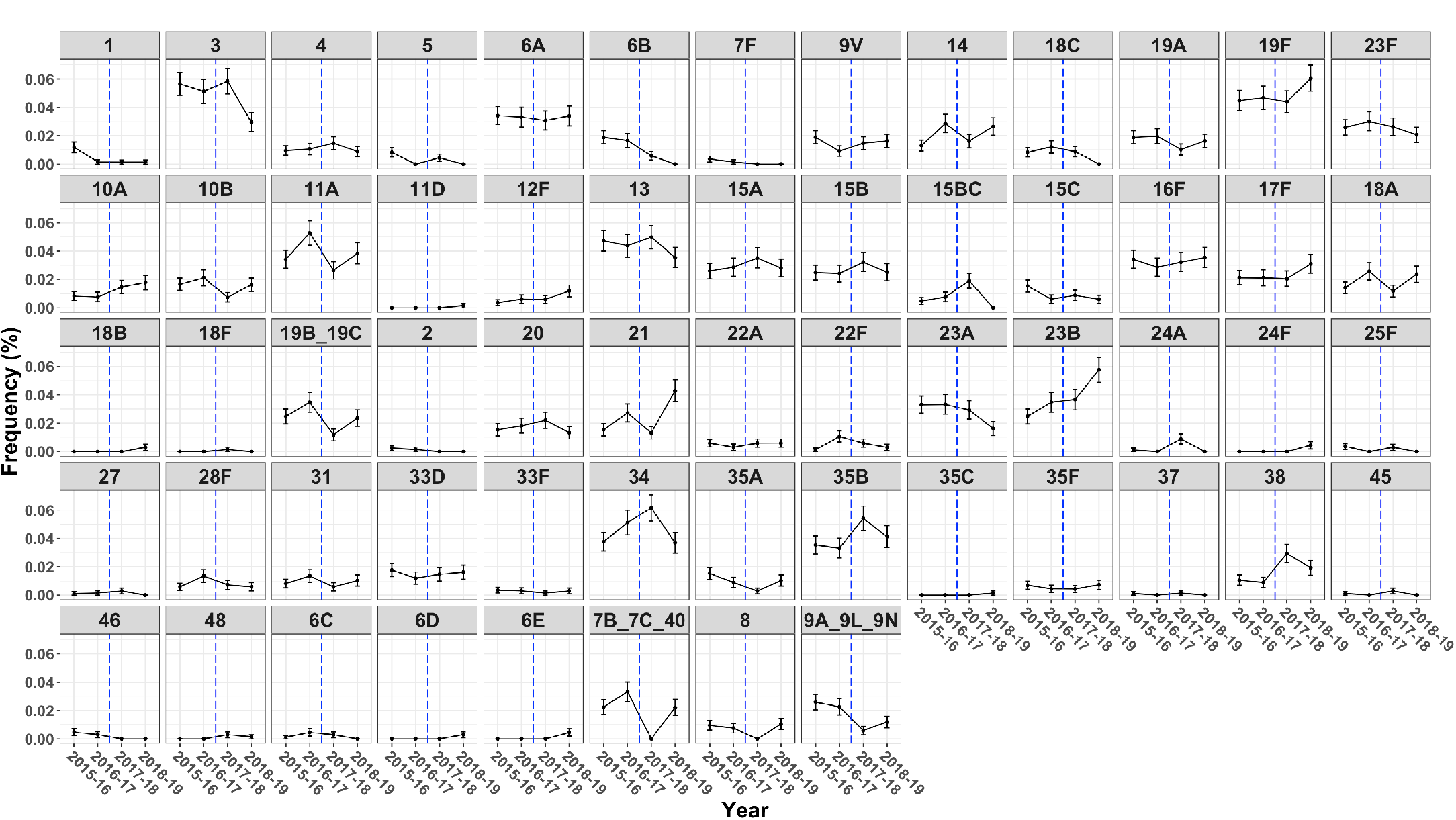
Frequency of *S. pneumoniae* serotypes during carriage surveillance between June 2015 and June 2019 (surveys 1-8). Data points refer to: 2015-16 = community carriage surveys 1 and 2, 2016-17 = surveys 3-4, 2017-18 = surveys 5-6, 2018-19 = surveys 7-8. The dashed blue line separates early-stages of carriage surveys (June 2015 to April 2017, survey 1-4) from late-stages of carriage surveys (May 2017 to June 2019, survey 5-8). The upper row shows vaccine serotypes (1 to 23F), the following rows show non-vaccine serotypes. Serogroups are determined according to Pneumocat results. Non-typeable strains are excluded from the analysis. Error bars show standard error of the mean.

### Serotype switching amongst both vaccine and non-vaccine serotypes

On the basis of the sequence-typing, we identified 44 separate serotype switching events in the Blantyre collection, described by a total of 22 STs which corresponded to more than one serotype (Table 2).

**Table 2.**
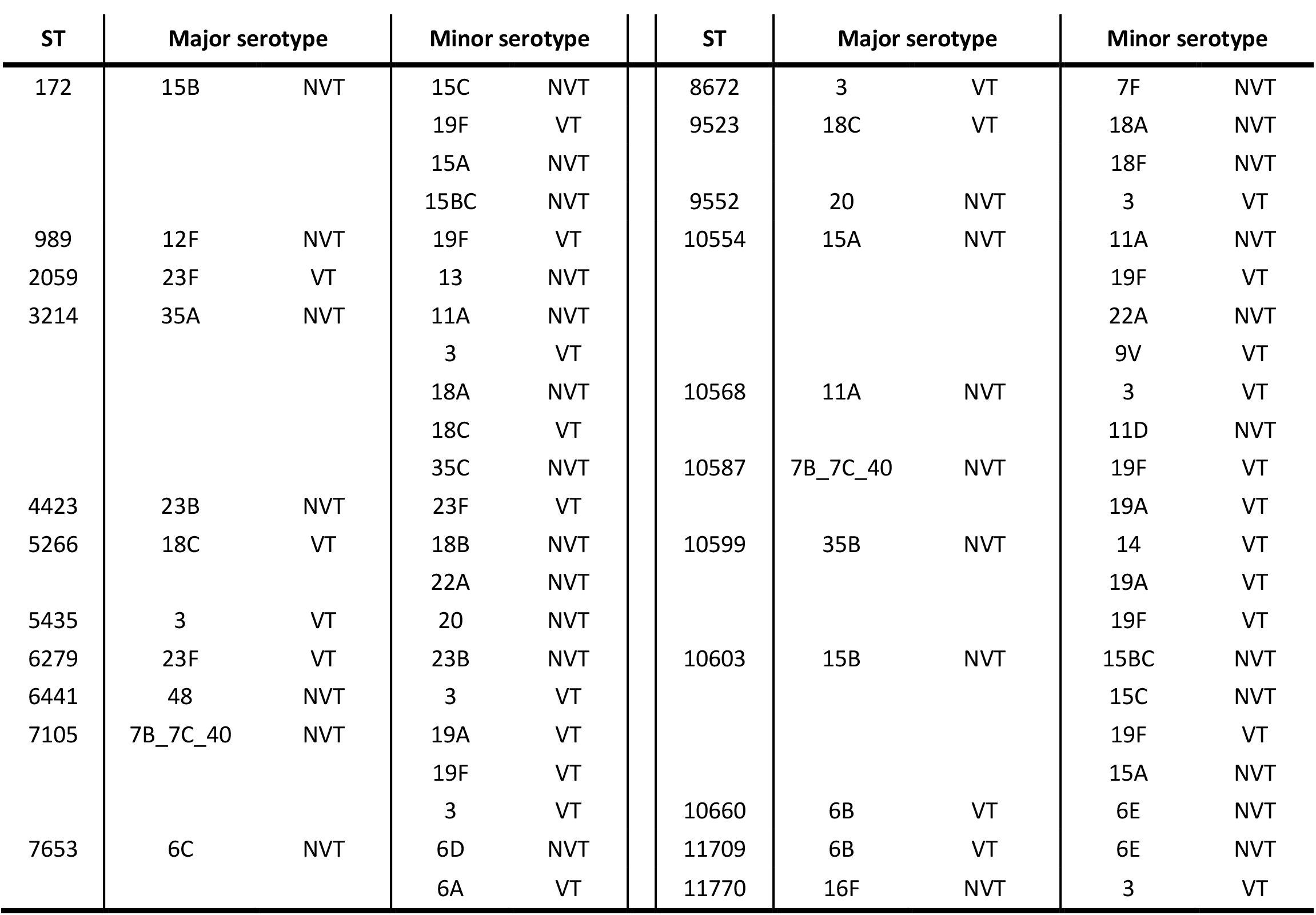
Capsular switch events. For each ST we present the most common serotype (major), the least common serotypes (minor serotype, described in the text as switch serotypes) and whether each serotype is included in PCV13 (VT and NVT).

| ST | Major serotype |  | Minor serotype |  |  | ST | Major serotype |  | Minor serotype |  |
| --- | --- | --- | --- | --- | --- | --- | --- | --- | --- | --- |
| 172 | 15B | NVT | 15C | NVT |  | 8672 | 3 | VT | 7F | NVT |
|  |  |  | 19F | VT |  | 9523 | 18C | VT | 18A | NVT |
|  |  |  | 15A | NVT |  |  |  |  | 18F | NVT |
|  |  |  | 15BC | NVT |  | 9552 | 20 | NVT | 3 | VT |
| 989 | 12F | NVT | 19F | VT |  | 10554 | 15A | NVT | 11A | NVT |
| 2059 | 23F | VT | 13 | NVT |  |  |  |  | 19F | VT |
| 3214 | 35A | NVT | 11A | NVT |  |  |  |  | 22A | NVT |
|  |  |  | 3 | VT |  |  |  |  | 9V | VT |
|  |  |  | 18A | NVT |  | 10568 | 11A | NVT | 3 | VT |
|  |  |  | 18C | VT |  |  |  |  | 11D | NVT |
|  |  |  | 35C | NVT |  | 10587 | 7B_7C_40 | NVT | 19F | VT |
| 4423 | 23B | NVT | 23F | VT |  |  |  |  | 19A | VT |
| 5266 | 18C | VT | 18B | NVT |  | 10599 | 35B | NVT | 14 | VT |
|  |  |  | 22A | NVT |  |  |  |  | 19A | VT |
| 5435 | 3 | VT | 20 | NVT |  |  |  |  | 19F | VT |
| 6279 | 23F | VT | 23B | NVT |  | 10603 | 15B | NVT | 15BC | NVT |
| 6441 | 48 | NVT | 3 | VT |  |  |  |  | 15C | NVT |
| 7105 | 7B_7C_40 | NVT | 19A | VT |  |  |  |  | 19F | VT |
|  |  |  | 19F | VT |  |  |  |  | 15A | NVT |
|  |  |  | 3 | VT |  | 10660 | 6B | VT | 6E | NVT |
| 7653 | 6C | NVT | 6D | NVT |  | 11709 | 6B | VT | 6E | NVT |
|  |  |  | 6A | VT |  | 11770 | 16F | NVT | 3 | VT |

**Table 2.** Example of genes specific of MTs that expanded at late stages of carriage survey identified via pan-GWAS. Columns show the annotation of the genes or the gene system, the MTs and serotypes in which the gene was found, the sensitivity and specificity related to its presence in the MT reported in the table (and its absence in the other MTs). Full table is shown in supplementary material (Table S2). All p-values (adjusted using a Bonferroni correction) were <0.01, as reported by Scoary.

|  | **MT** | **Serotype** | **Sens./Spec.** |
| --- | --- | --- | --- |
| PTS system mannose-specific | 120 | 38 | 100/100 |
| PTS system sorbose-specific | 120 | 38 | 100/100 |
| PTS system galactitol-specific | 93 | 23B | 100/100 |
| PTS system lactose-specific | 93 | 23B | 100/100 |
| TA system pezA-pezT | 120 | 38 | 100/100 |
| TA system yefM-yoeB | 93/120 | 23B/38 | 100/100 |
| Competence-stimulating peptide type 1 - CSP1 | 120 | 38 | 100/100 |
| Tetracycline resistance - tetM | 120 | 38 | 95.6/100 |
| Multidrug resistance ABC transporter - bmrA | 93 | 23B | 100/98.07 |
| Lactococcin-G-processing and transport - lagD | 109/74 | 17F/10A | 93.1/92.1 |
| Bacteriocin lactacin-F - lafA | 109 | 17F | 100/100 |
| Alpha-hemolysin translocation ABP - HlyB | 109/74 | 38/10A | 100/100 |
| Protease – ydcP | 33 | 34 | 98.8/97.9 |

Directionality of these switch events was determined by considering the genetic relatedness of the strains and assuming that the most common serotype (capsule type in a dominant lineage) within an ST was the serotype from which the switch originated [Chaguza, *et al*., 2017]. Rather than a clear shift from VTs towards NVTs, low-frequency VT or NVT variants were identified to undergo switching throughout the carriage surveys in several ST and GPSC lineages (Figure S2a and S2b). These data support the emerging evidence that even in the context of vaccine introduction, the process of serotype switching is not limited to VTs (e.g. ST7653 6C/6D or ST3214 35A/11A). We suggest that although a serotype switch may be directly vaccine-induced, switched serotypes may have occurred stochastically and then emerged alongside other vaccine-associated changes in population structure. Indeed, it seems likely that selection forces that are more complex than serotype-directed vaccine pressure alone facilitate many of such switches such as environmental and antimicrobial pressures [Croucher, *et al*., 2015].

### Shifts in metabolic gene profiles of the pneumococcal population

To further identify dynamic changes in the pneumococcal population structure beyond serotype switching, we defined metabolic genotypes (*metabolic types, MTs*) based on our previously described theoretical framework [Watkins, *et al*., 2015]. Each MT was classified on the basis of differences in core genes involved in metabolism and energy production, following functional annotation of the pan-genome of the pneumococcal population. This typing method therefore relies on sequence differences between genes involved in bacterial metabolism (i.e. function), rather than lineages defined exclusively by bacterial evolutionary history.

We grouped the Blantyre pneumococcal carriage isolates into 148 discrete MTs (Figure 2). As shown in the clustering tree, these MTs were neither independent of serotype nor did they completely overlap with serotype. For instance, the 14 different serotype 19F MTs were spread across the clustering tree. Overall, the majority of MTs did not significantly change in frequency when comparing early and later surveillance isolates (p>0.05; Fisher’s exact test), highlighting a high residual diversity within the bacterial population (figure S3a and S3b). Nonetheless, our analysis highlighted a shift in the pattern of several MTs: for instance, with the NVTs 10A, 17F, 34, 38 and 23B which showed a significant increase in MTs 74, 109, 33, 120 and 93 (Figure S3a and S3b, p<0.05, Fisher’s exact test).

**Figure 2.**
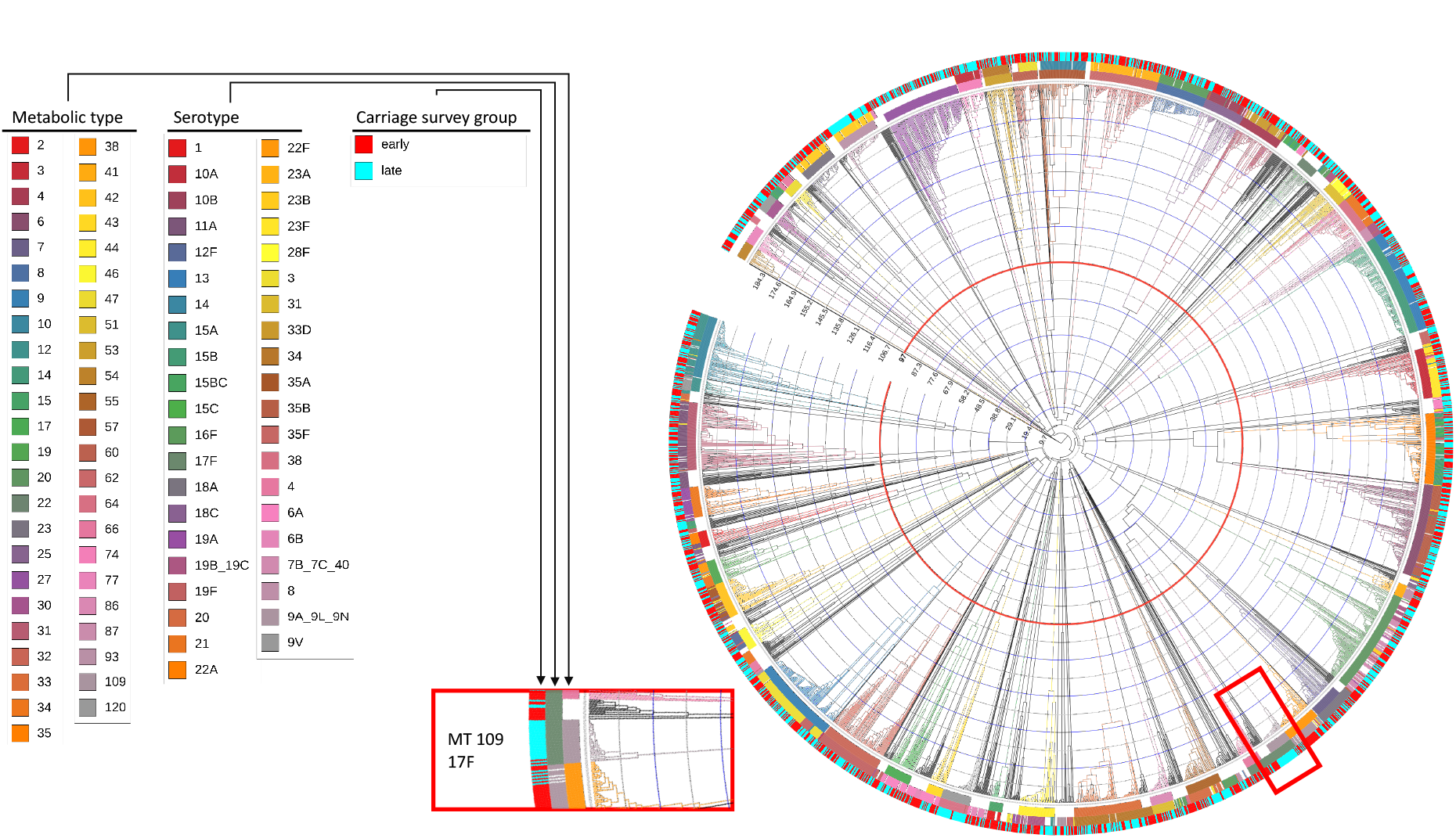
Hierarchical clustering dendrogram of *S. pneumoniae* metabolic types amongst carriage isolates (June 2015 - June 2019). The dendrogram is based on of the hamming distance between the different allelic profiles of the metabolic genes. Coloured strips represent the Metabolic type (also reported by an exterior number annotating the branch), the serotype, and the carriage survey group (early = survey 1 to 4, late = survey 5 to 8). MTs are defined by curtailing the dendrogram at half the maximum distance, indicated by the red line (n=148 discrete MTs, containing only MTs with more than 15 observations). The inset show a magnification of the dendrogram around serotype 17F/MT109.

In total we observed 32 instances in which VTs switch capsular serotype to NVT, or vice versa, while maintaining the same metabolic profile which is in line with a vaccine-induced metabolic shift as described in our previous models [Watkins, *et al*., 2015]. However, we cannot infer vaccine causality from this dataset.

### Metabolic genotypes increasing in frequency are characterised by increased AMR

To address the hypothesis that vaccination drives changes in the pattern of AMR by removing the competition of non-AMR VTs [Obolski, *et al*., 2018], we assessed whether the emerging MTs amongst the NVTs 10A, 17F, 23B, 34 and 38 were also associated with increased AMR. We determined resistance genetically to penicillin, an important first-line antibiotic for pneumococcal disease, together with chloramphenicol, erythromycin and tetracycline, which are also commonly used antibiotics in this population. Higher penicillin MICs (minimum inhibitory concentration) were observed in the later surveillance period compared to the early surveillance period (Wilcoxon rank sum test, p<0.01) (Supplementary Figure S3e). The majority of the serotypes and MTs identified in this dataset (n=39 and n=79 respectively) were significantly associated with either a positive or negative association with penicillin AMR (i.e. MIC > 0.06 ug/ml, p<0.05; Fisher’s exact test), indicating that this phenotypic characteristic is likely linked to the isolate’s genotype [Chaguza, *et al.,* 2019] (Supplementary Figure S3c and S3d).

We observed several instances in which emerging metabolic genotypes were associated with higher AMR. Among NVTs 17F, 10A, 23B, 34 and 38 we show that MTs 109, 74, 93, 22 and 120 (respectively, Figure 2) were more frequently identified in the later surveillance period (p<0.05; Fisher’s exact test) (Figure 3), indicating a shift in the dominant genotype observed within each serotype. In four of these NVTs (17F, 10A, 23B and 38), the emergence of MTs in later surveillance was associated with increased AMR: either in terms of an increased MIC for penicillin (genetically assessed, Figure 3) or in terms of presence of other antibiotic resistance genes such as *tetM* and *ermB*; Supplementary (Figure S4). In serotypes 17F, 23B and 38, there was an increase in the frequencies of MT114, MT97 and MT122, respectively. In each case, the MT that emerged showed a higher penicillin MIC, compared to MTs common in the bacterial population at earlier surveys stages. Interestingly, the increased frequency of MT46 of serotype 34 in later surveillance surveys without exhibiting an increase in AMR suggests a variety of forces that drive the selection and restructuring of bacterial populations.

**Figure 3.**
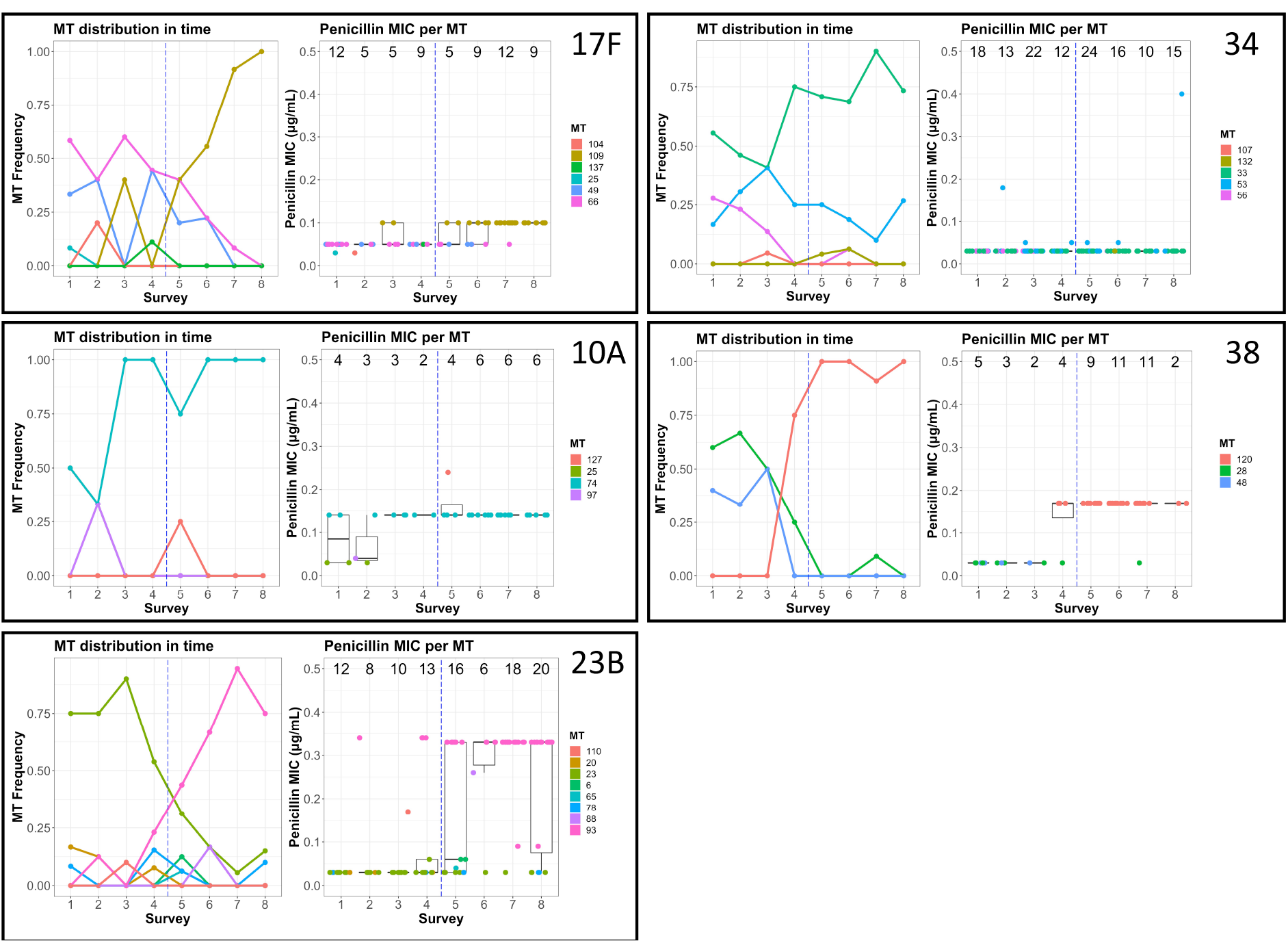
Shifts in metabolic profiles and penicillin MIC over time (surveys 1-8; June 2015 – June 2019). Boxes show the frequency of isolation of each metabolic profile per survey (“MT distribution in time”) and penicillin MIC of each isolate (box and whiskers plot, with points representing isolates, and numbers on top the number of isolates in each survey) for to serotypes 17F, 10A, 23B, 34 and 38. Vertical blue lines separate the early-late survey periods. Presence of AMR genes per serotype is shown in Figure S4.

There were no significant changes (p>0.05, Fisher’s exact test) in carriage frequency of the majority of the VTs over the duration of the surveillance period - i.e. they did not increase nor decrease when comparing the early and later periods of surveillance. These included, 3, 19F and 23F (Figures 1 and S3). Serotypes 3 and 23F in particular were found at relatively high frequencies after vaccine introduction in other settings [Kandasamy, *et al*., 2019; Lewnard and Hanage, 2019]. Contrary to what was described with NVTs, the persistent VTs in this dataset were characterised by the presence of a dominant MT (MT 9 for serotype 3, MT 3 for serotype 23F) consistently present during the survey period, showing increased penicillin MIC and the presence of antimicrobial resistance genes (Figure S5).

Within those serotypes where we identified genotype switch events (namely, 17F, 10A, 23B, 38 and 23B), we investigated genetic relatedness amongst MTs and time-calibrated phylogeny, in order to reconstruct the emergence of those recent dominant MTs. We assessed the genetic relatedness between MTs, hence their likelihood to have recently expanded clonally, by calculating the core genome SNP (single nucleotide polymorphism) divergence between MTs (Figure S6). For serotypes 38, 17F and 34, recently identified MTs were characterized by lower genotypic difference (lower SNP variance, p<0.05, F-test for variances, Figure S6), indicating that these isolates expanded clonally. This recent clonal expansion was confirmed by the temporal signal identified in the phylogeny (Figure S7). Root-to-tip regression highlighted that the emergence of the recently identified lineages (MT109 and MT120) in serotype 17F and 38 was likely to have happened before the year 2011 (17F MT114: 2000.9 [95% CI 1988.35 - 2008.1] node 95, 2009.7 [2005.7 – 2012.3] node 94; for 38 MT122: 2009.3 [2000.3 – 2013.3] node 85, 2007.5 [1996.7 – 2012.2] node 84, 2004.1 [1985.2 – 2011.4] node 83). Thus, none of these MTs are likely to have originated after PCV13 introduction, but rather we propose that they existed as variants at low frequency before vaccine introduction that underwent clonal expansion. It was not possible to temporally calibrate the phylogeny of serotypes 23B, 10A and 34, hence we could not infer the time of emergence of those lineages. This may be due to the smaller sample size of some lineages (i.e. in serotype 10A) or to lineages that originated much earlier than their isolation in this study.

### Emerging metabolic genotypes show multiple adaptations

Having demonstrated a shift in MTs in serotypes 10A, 17F, 23B, 34 and 38 associated with increased AMR during the post-vaccine introduction observation period, we then assessed whether these emerging MTs had associated virulence characteristics that could convey a competitive advantage. To do this, we investigated the accessory genome using a pan-genome wide association study approach (pan-GWAS, Gori, *et al*., 2020). We identified 93, 73, 51, 30 and 175 genes present exclusively in the most common emerging metabolic profiles of serotypes 10A, 17F, 23B, 34 and 38, respectively (Figure S8; full gene list is reported in Supplementary Table S1; examples of genes associated to each MT analysed are shown in Table 2). Functional annotation showed that these included genes responsible for metabolism and carbohydrate transport. In serotype 38 MT120, for instance, mannose- and sorbose-specific transporter genes, absent in other MTs, were identified. MT93 serotype 23B was characterised by lactose and galactiol transporters (Table 2) which were absent in other MTs. The emerging MTs were also characterised by the presence of bacterial defense systems, such as multidrug resistance ABC transporters, restriction enzymes and antimicrobial resistance genes (Table 2).

The MT93 and MT120 of serotypes 23B and 38 accessory genomes encode several Toxin-Antitoxin (TA) systems. Variants of the *pezAT* system was present in MT93 and MT120 and a *yefM*-*yoeB* system was present in MT120. This particular TA-system is located in a previously described integrative-conjugative region (ICE) [Brown *et al.,* 2003], which is absent from the non-MT120 strains. The ICE region is present in several non-pneumococcal *Streptococci*, such as *dysgalactiae* and *suis* [Huang, *et al.,* 2016], suggesting that its acquisition may have occurred via horizontal gene transfer (HGT). This ICE also carries specific metal transporters, phage resistance and competence systems, as well as a previously described agglutinin receptor involved in colonisation [Brown, *et al*., 2003].

Together, these data demonstrate that the emerging MTs have acquired a range of potential adaptions (Figure 4). In populations with a high force of infection and host vulnerabilities such as malnutrition and HIV, this potential for competitive advantage in nutrient transport, cellular metabolism, nasopharyngeal epithelial colonisation and AMR could lead to a resurgence in invasive NVT pneumococcal disease.

**Figure 4.**
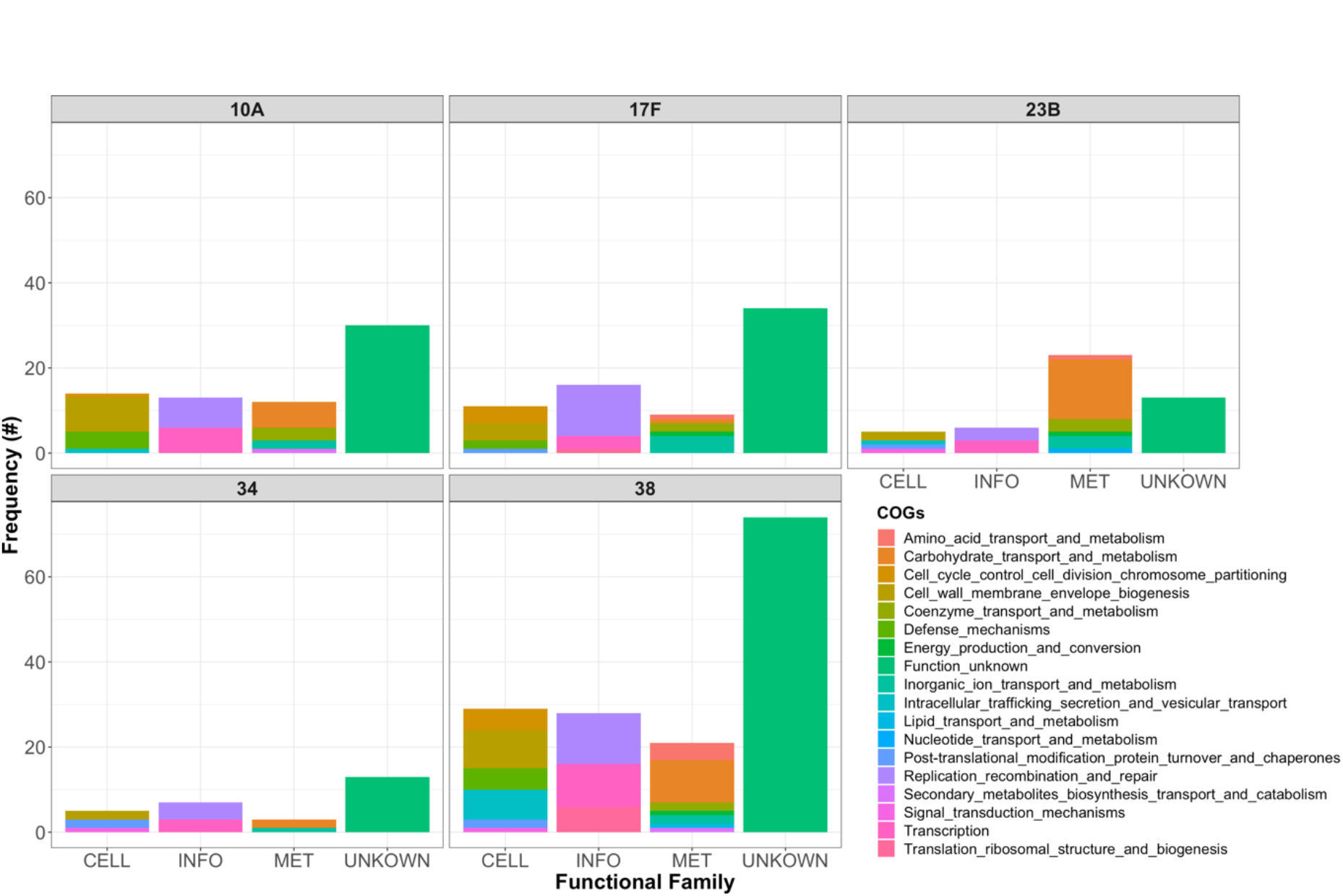
Clusters of orthologous genes enriched in Serotype 10A (MT), 17F (MT), 23B (MT), 34 (MT), 38 (MT). Each barplot shows the number of genes in each functional class for the dominant metabolic type in the 5 serotypes, as reported by the pan-GWAS analysis. The columns refer to the functional classes “Metabolism” (MET), “Cellular processes and signaling” (CELL), “Information storage and processing” (INFO), or poorly characterized genes (UNKNOWN).

### Phenotypic characterisation of emerging MT reveals adaptations that may confer competitive advantage

To further explore this potential competitive advantage amongst emerging NVT metabolic genotypes with increased AMR, we compared MT shifts within serotype 38 and serotype 23B in several *in vitro* phenotypic assays. These aimed at representing some of the challenges of bacterial lifestyle in the nasopharyngeal niche. Firstly, we cultured combinations of different MTs in standard growth medium [Ram, *et al.,* 2019]. The growth parameters and the *in-vitro* fitness were calculated by *Curveball* for MTs 28 and 120 for serotype 38 and MTs 23 and 93 for serotype 23B (Figure 5). This software fits the growth curves of the different bacterial strains to several growth models and attempts to predict which one would have a competitive advantage over the other while sharing resources.

**Figure 5.**
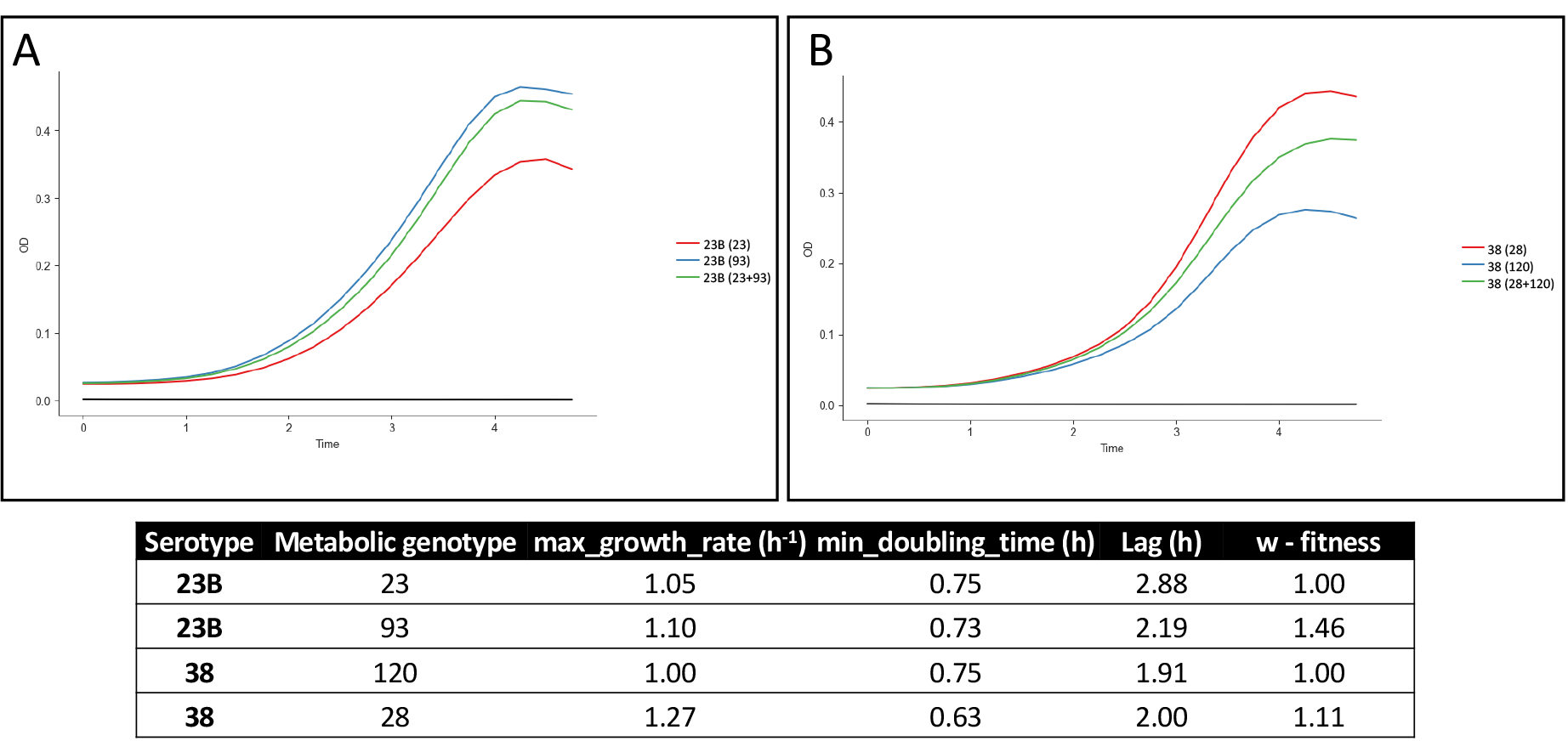
Growth curves and inferred in-vitro fitness of different metabolic types in serotype 23B and serotype 38. The growth curves of the MTs comprising serotype 23B (a) and 38 (b) separately (blue and red curves) and in an *in-vitro* co-culture (green curve). For each curve the MT or the combination of MTs is reported in parentheses. The table shows the growth parameters as predicted by Curveball (nu - deceleration parameter, q0 - initial physiological state, r - specific growth rate in low density [Ram *et al.,* 2019].

Within serotype 23B MT93, the emergent strain reached a higher maximum optical density (OD) than MT23 (Figure 5). Using the Baranyi-Roberts model [Ram, *et al*., 2019] to approximate co-culture growth, we found that MT93 is expected to have a higher *in-vitro* fitness than MT24. Within serotype 38 MT120, the emergent strain, grew at a lower rate and to a lower maximum OD than MT28 (Figure 5) and therefore had a lower predicted fitness than MT28. Thus, while some emergent MTs have a growth advantage, others do not.

The epithelial colonisation process is a prerequisite for both disease and onward transmission to another host [Weiser *et al*., 2018]. We therefore then tested these MT pairs in a well-standardised Detroit 562 human nasopharyngeal epithelial infection model [Weight, *et al*., 2019]. Within serotype 38, we observed no difference in the ability to colonise the epithelial cells between MTs 28 and 120. In contrast, serotype 23B, MT93 was internalised into epithelial cells at a higher frequency than MT23 (Figure 6a and 6b) even though both genotypes were characterised by the same ability to associate with epithelial cells. We have previously suggested that differences in this so-called epithelial “microinvasion” may be fundamental to the outcome of colonisation, explaining strain differences in the potential of *S. pneumoniae* to cause invasive disease, persist during carriage and be transmitted from person-to-person [Weight, *et al*., 2019].

**Figure 6.**
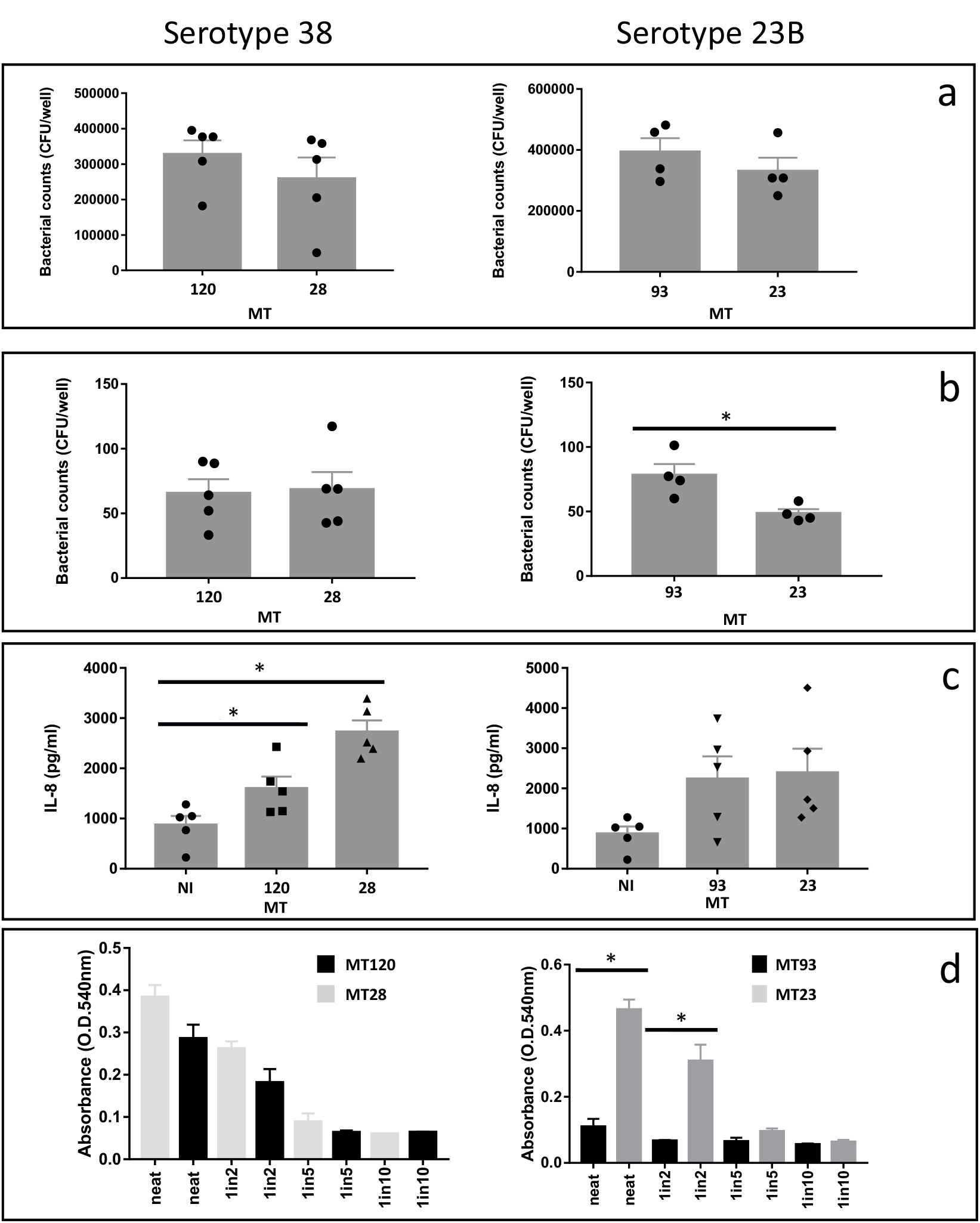
Adhesion and invasion to epithelial cells, IL-8 secretion and haemolysis for MT122 and 29 (serotype 38) and MT91 and 24 (serotype 23B). (a) Number of bacteria adhered to epithelial cells, recovered after 2 hours of incubation, (b) Number of bacteria invading epithelial cells after 2 hours incubation, (c) IL-8 produced by epithelial cells, incubated with bacterial cells for 6 hours. NI = non-infected control. (d) haemolysis promoted by the different bacterial strains (lower OD corresponds to higher haemolysis), for different bacterial dilutions (neat ∼ 1.5×10^6^ bacterial cells. Results for 1/2, 1/5 and 1/10 dilutions are shown). Each experiment was repeated at least 3 times independently. In a, b, and c, dots on the bar plots show the result of each repetition. (*= p<0.05. (a), (b), (d) : T-test; (c) one-way ANOVA).

Other phenotypic characteristics in different MTs were assessed, such as their ability to stimulate the production of the phagocyte chemoattratctant IL-8 in epithelial cells (Figure 6c) and their haemolytic potential, a measure of functional pneumolysin production (Figure 6e). Serotype 38’s MT120 stimulated lower IL-8 production, compared to MT28, while the two MTs in serotype 23B did not stimulate a higher IL-8 production when compared to the non-infected control. The haemolytic potential of serotype 23B’s MT 23 was higher than serotype 23B’s MT93 (Figure 6e), while there was no significant difference was identified between serotype 38 strains.

Together these data suggest that the emerging MTs have not arrived at a single adaptive profile but have undergone a variety of changes (growth; epithelial invasion; inflammation; and pneumolysin production) that may have enabled them to become more prominent in the context of vaccine introduction.

## Discussion

How a pneumococcal population evolves both under natural or vaccine-induced pressures remains an open question. It has been long known that *S. pneumoniae* is characterised by a high level of genome plasticity, and this species diverges more quickly than many other mucosal bacterial species associated with vaccine preventable disease [Tettelin and Hollingshead, 2004; Croucher *et al.,* 2011]. Here, using a large post-PCV introduction pneumococcal carriage surveillance dataset from an urban population in Malawi, we describe the emergence of pneumococcal lineages with unique virulence, AMR and metabolic characteristics. Rather than using the populational structural methods of the Pneumococcal Molecular Epidemiology Network (PMEN) [McGee, *et al*., 2001] or the Global Pneumococcal Sequencing (GPS) consortium [Gladstone, *et al* 2019], our genotyping approach defined pneumococcal lineages based on metabolic genotypes (MT) [Watkins, *et al*., 2015].

We show that in addition to serotype replacement, there has been a shift in the profile of AMR MTs among NVTs characterised by exclusive genes responsible for metabolism and carbohydrate transport, and toxin-antitoxin systems located in an integrative-conjugative region suggestive of horizontal gene transfer. Phenotypically, these emergent genotypes were found to have differential growth, haemolytic, inflammatory or epithelial adhesion/invasion traits that may confer advantage in the nasopharyngeal niche. These findings support theoretical models that predict that PCV introduction disrupts complex competitive interactions between colonising pneumococci, with the potential to promote replacement with fitter, more virulent, and more antimicrobial resistant existing pneumococcal lineages [Watkins, *et al*., 2015; Obolski, *et al*. 2018]. We postulate that if these emergent NVT MTs become fixed in the population, they may lead to a lower vaccine effectiveness and vaccine escape. We also observed stable MTs among VTs and although our observations are limited to the post-PCV13 era, we speculate that these genotypes may confer a competitive advantage leading to inadequate PCV13 effectiveness in the control of VT carriage, including serotypes 3, 19F and 23F.

The overall effect of the shift in MTs that we have observed is dependent on their transmissibility and their relative invasiveness [Lo, *et al*., 2019]. These MTs have emerged within serotypes that are known to be important contributors to IPD and are commonly carried. Serotype 38 has high heterogeneity in its invasive potential in South Africa and the USA [Lo, *et al*., 2019]; serotype 23F ST2059/MT3 was reported to have a higher risk for invasiveness in strains isolated in South Africa and in the USA; serotype 23B ST4423/MT97 was identified in IPD in South Africa and Israel; and serotype 35B, which also expanded in frequency in this study, was highlighted as an expansion serotype in Malawi IPD [Lo, *et al*., 2019; Gladstone, *et al*., 2019]. Moreover, serotypes 23B and 38 are among the top 10% of serotypes causing IPD after vaccine introduction in Europe, Asia and US [Balsells, *et al*., 2017].

The emergence and expansion of particular NVTs in the period after PCV introduction has been widely reported [Lo, *et al*., 2019], and the serotype switching processes and dynamics highlighted here have also been reported by others [Chaguza, *et al*., 2019; Gladstone, *et al*., 2019]. For instance, the serotype switch event involving ST8672 (serotypes 3 and 7F) was also identified in a post-vaccination carriage study in northern Malawi [Chaguza, *et al*., 2019]. Similarly, the Global Pneumococcal Sequencing study, identified ST10599 and ST989 as part of switch events, highlighting how these mechanisms are common in pneumococcal genotype and serotype dynamics [Lo, *et al*., 2019].

To assess the generalisability of our findings, we undertook a wider comparison using pneumococcal genomes sequences from other African countries [Lo, *et al*., 2019; Gladstone, *et al*., 2019]. This revealed the presence of analogous MTs outside Malawi, leading to the hypothesis that these MTs have become more prevalent across the continent following PCV introduction. In particular, MT97 and MT122 (associated with serotypes 23B and 38 respectively) have been identified after vaccine introduction in invasive pneumococcal disease isolates in both South Africa and The Gambia [Gladstone, *et al.,* 2019]. However, interestingly, we found that rather than originating from a common source, differences in the core genome SNPs among MT122 strains isolated in South Africa and in Malawi (∼6000 SNPs, Figure S8) suggest that these different lineages originate from different ancestors that have subsequently converged to the same MT.

A theme that has emerged from our analysis is that although the emergent MTs have successful metabolic, virulence and AMR properties that may confer a competitive advantage over other MTs, we did not identify genetic or phenotypic traits that suggest a single adaptive profile amongst the emergent MTs. In contrast, we identified multiple different adaptations that may provide an advantage in distinct ecological niches. With regards to the accessory genome, for instance, some MTs are characterised by particular sugar transporters, TA-systems and bacteriocins production systems, which would confer competitive advantage while sharing resources. In particular, a characteristic of the MTs that appeared to have emerged in recent years in our study population is the presence of genes that encode particular toxin/antitoxin systems. These systems are present in a variety of bacterial species both at a plasmid and chromosomal level, and involved in a variety of cellular processes – including virulence, environmental stress response and niche adaptation [Van Melderen and De Bast, 2009; Page and Peti, 2016; De La Cruz, *et al*., 2013; Butt, *et al*., 2014; Kim, *et al*., 2009; Wen, *et al*., 2014]. The pan-GWAS analysis highlights the presence of virulence factors in expanding MTs: in MT33 for instance the presence of a *ydcP* protease, a collagenase which facilitates breaking of extracellular structures and a known virulence factor in other bacterial species [Navais *et al.,* 2014], identified to be associated with pneumococcal IPD [Obolski, *et al*. 2019]; and in MT109 and 74 of a hemolysin (pneumolysin) translocation ATP-binding protein gene, known virulence factors for *S. pneumoniae*. Together, these observations indicate that the expanding metabolic types accumulate characteristics that may affect niche competition and virulence.

Recent studies have described bacterial fitness in different terms, adapting its definition to the available data. For instance, recent studies describe the negative frequency dependent selection in *S. pneumoniae* populations, whereby bacterial fitness is affected by the frequency of rare accessory genes [Corander, *et al*., 2017; Azarian, *et al*.; 2020]. We have used growth curve analysis to infer relative, mixed-culture bacterial fitness [Ram, *et al*., 2018]. In combination with previous modelling analyses, the results described in this work is consistent with a complex and diverse behaviour of a *S. pneumoniae* population under vaccine pressure: serotype 23B MTs for instance, behaves closely to the VIMS model description whereby the fitter genotype which was suppressed beforehand expands after vaccine introduction due to niche clearance (Figure 3). On the other hand, serotype 38’s MTs behave in accordance with the hypothesis of an increase in frequency of AMR NVTs, driven by vaccination [Obolski *et al*., 2019]. An antibiotic susceptible MT of serotype 38 exhibited a higher intrinsic fitness, as determined by the growth curve analysis. However, it appears that this MT was overtaken by another, antibiotic resistant MT of serotype 38, when vaccination alleviated the competition pressure from VTs.

We also found that the most common strain of 23B (MT93) is less haemolytic then MT23 and more successful in invading nasopharyngeal cells. On the other hand, serotype 38 MT120 stimulated less IL-8 production, when compared to a less common metabolic genotype (MT28). Pro-inflammatory interleukin-8 attracts neutrophils to the site of infection [Bickel, 1993] and therefore by evading the immune system MT120 may have a competitive advantage.

The genetic and phenotypic analyses described here highlighted that the way the single strain evade the immune system, or is more successful in transmitting or colonising different ecological niches, is multifactorial. We cannot describe a common phenotype characteristic between all the genotypes that have expanded recently, but we highlighted several differences that could help the success of each genotype in different situations [Watkins, *et al*., 2015].

The main limitation of this analysis is the lack of genomic data prior to the introduction of PCV in Blantyre. However, we have been able to leverage this large dataset to capture ongoing changes in a population with high vaccine coverage [Swarthout, *et al.,* 2020]. Because of the cross-sectional nature of the surveillance contributing to the Blantyre collection we were not able to track the emergence of MTs within individual hosts over time. Finally, as is inherent in observational studies, we have not been able to attribute causality to routine vaccine introduction.

In conclusion, this study highlights that in this high burden vaccine-exposed population, AMR and a range of genotypic and phenotypic traits facilitate the remarkably rapid expansion of MTs in the community. Vaccine strategies that increase PCV valency may be insufficient to interrupt this process [Colijn, *et al*., 2020]. The multiple adaptations by these MTs extend beyond simple serotype replacement, and successful adaptation could further undermine vaccine control and promote the spread of AMR.

## Methods

### Bacterial Isolates and Carriage Surveys

The strains analysed in this study were isolated as part of a prospective observational study using stratified random sampling to assess pneumococcal nasopharyngeal carriage in Blantyre, Malawi [Swarthout, at al., 2020]. In brief, sampling consisted of twice-annual rolling cross-sectional surveys over the course of 4 years amongst PCV13-vaccinated children 2-7 years old, PCV13-unvaccinated children 5-10 years old, and HIV-infected adults 18-40 years old on ART. Isolates from the HIV-infected adults were sequenced from surveys 1, 2, 5 and 6 only.

### DNA Sequencing and Assembly

NP swabs were stored in skim-milk-tryptone-glucose-glycerol (STGG) at -80C. After being thawed and vortexed, 30 µL of STGG was plated on gentamicin-sheep blood agar (SBG; 7% sheep blood agar, 5 µL gentamicin/mL) and incubated overnight at 37 °C in 5% CO2. Plates showing no *S. pneumoniae* growth were incubated overnight a second time before being reported as negative. *S. pneumoniae* was identified by colony morphology and optochin disc (Oxoid, Basingstoke, UK) susceptibility. The bile solubility test was used on isolates with no or intermediate optochin susceptibility (zone diameter < 14 mm). A single colony of confirmed pneumococcus was selected and grown on a new SBG plate as before. Growth from this second plate was used for serotyping by latex agglutination (ImmuLex™ 7-10-13-valent Pneumotest; Statens Serum Institute, Denmark) [Swarthout *et al* 2020]. DNA was extracted from an overnight lawn plate culture from isolates archived after serotyping, using DNAeasy blood and tissue kit (Qiagen) following the manufacturer’s guidelines and sequenced using HiSeq4000 (paired-end library 2 × 150) platform at Oxford Genomics Centre UK.

Raw DNA reads were trimmed of low-quality ends and cleaned of adapters using Trimmomatic software (ver. 0.32). De novo assembly was performed with SPAdes software (ver 3.8.0), using a sample of 1,400,000 reads and k-mer values of 21, 33, 55, and 77. De novo assemblies were checked for plausible length (between 1,900,000 and 2,200,000 bp), annotated using Prokka (ver. 1.12), and checked for low-level contamination using Kraken (ver. 0.10.5). In cases for which more than 5% of the contigs belonged to a species different from *Streptococcus pneumoniae*, the genome sequence was not included in any further analysis. Resulting assemblies are available on pubmlst.org (https://pubmlst.org/organisms/streptococcus-pneumoniae isolates numbers are in table S2). Metadata of the strains used in this work are reported in table S2.

### Definition of Serotype, multilocus sequence type, global pneumococcal sequencing cluster and AMR

Serotypes were determined via DNA sequence, using the PneumoCaT software (Ver. 1.2.1, https://github.com/phe-bioinformatics/PneumoCaT). Phenotypic serotype definition by latex agglutination was used as validation of the genetic serotype definition, observing a correlation of over 90%, in alignment with previous estimations [Swarthout, *et al*., 2021]. For the analyses in this work, serotype determined by DNA sequencing was used.

Multilocus sequence types (MLST, STs) were assigned using the databases from the allelic profiles of 7 housekeeping genes (*adhP, pheS, atr, glnA, sdhA, glcK*, and *tkt*). BLASTn was used to align the DNA assemblies to the DNA fragments typical of each allele for each house keeping gene (parameters: E value 1e−10, minimum 95% identity, minimum 90% query coverage). This grouped strains into 200 unique STs. Strains which did not show a full set of housekeeping gene alleles or were not assigned to any previously described ST (n = 783) were double-checked for sequence contamination and assigned to a new sequence type definition. The database of MLST housekeeping fragments was downloaded from pubmlst.com/Spneumoniae in October 2019. The GPSC genetic typing method (Global Pneumococcal Sequence Clusters) was described in several recent publications [Lo, *et al.,* 2019; Chaguza, *et al.,* 2018; Gladstone, *et al.,* 2019]. We collected the dataset to assign the GPSCs from https://www.pneumogen.net/gps/assigningGPSCs.html and used PopPUNK [Lees, *et al*. 2019] to cluster the isolates in the pre-defined GPSCs as described in Gladstone *et al*., 2019. AMR gene presence (for genes *cat, tetM, ermB, and mefA*) was detected via nucleotide-BLAST (E-value < 0.001, sequence coverage and identity >80%). For penicillin resistance, MIC was genetically assessed using the *pbp* genes allelic profile [Metcalf, *et al.,* 2016].

### Pangenome, definition of metabolic profiles and phylogenetic analyses

In order to capture the total variability of the population, a pangenome was generated from the combined isolates from surveys 1 to 8 using Roary (ver. 3.8.0). Parameters for each run were: 90% of minimum BLASTp identity; MLC inflation value 1.5; with 99% of strains in which a gene must be present to be considered “core”. Saturation of the pangenome was assessed. The core genome of this bacterial population included 1061 genes which was consistent with previous estimates from Malawi [Kulohoma, *et al.,* 2015]. The metabolic genes were selected via functional annotation of the core genome (n=386 core metabolic genes, Watkins, *et al*., 2015) through the eggNOG database (http://eggnogdb.embl.de). Functional classes selected to define the core metabolic genome were: D - Energy production and conversion, E - Amino acid transport and metabolism, F - Nucleotide transport and metabolism, G - Carbohydrate transport and metabolism, H - Coenzyme transport and metabolism, I - Lipid transport and metabolism, P - Inorganic ion transport and metabolism, Q - Secondary metabolites biosynthesis, transport, and catabolism.

Metabolic profiles were defined using the following pipeline: (i) for each genome the allelic profile of the metabolic core genes was defined using the Genome Comparator module of Bigsdb (bigsdb.com) [Jolley, *et al.,* 2018]; (ii) the hamming distance between each allelic profile was calculated (*hamming.dist* function of base R); (iii) the hamming distance between each isolate was used hierarchically cluster the population (*hclust*); (iv) the hierarchical clustering dendrogram was cut at midpoint to define the discrete metabolic profiles.

Maximum-Likelihood, recombination censored phylogeny was calculated with ClonalframeML, under a generalized time-reversible model with 100 bootstrap replicates. Core genomes used to produce ML phylogeny were calculated separately for the isolates belonging to the serotypes of interest (17F, 10A, 38, 34 and 23B) with Roary as described above, SNPsites (Ver 1.04) was used to identify the variable part of the core genome alignment which was then used for phylogeny reconstruction, Root-to-tip regression and phylogeny dating was calculated with BactDating R package (“xavierdidelot/BactDating”, 700000 permutations).

The pan-GWAS analyses were carried out using Scoary (https://github.com/AdmiralenOla/Scoary) as described in Gori, *et al*., 2020.

### Co-culture fitness and epithelial association/ invasion experiments

Strains BVY5TE, BVY11B (serotype 23B, MT 23 and 93 respectively), BVY2DJ and BVY123 (serotype 38, MT 120 and 28 respectively) were selected for phenotypic evaluation (Table S1).

*In-vitro* broth culture was carried out in order to apply the pipeline described by Ram *et al.,* in 2019. Isolates belonging to different MTs were harvested from a THY-10% glycerol stock and grown overnight on a Columbia agar plate with 5% horse blood (CBA) plate. The plate was used to seed 10 ml of Todd-Hewitt broth with 2.5% yeast extract (THY) to an optical density at 600nm (OD_600_) of 0.5-1, which was then diluted to an OD_600_ 0.05 and used directly as starter in at least 16 200ul wells of a 96-well plate. Each experiment also contained a mixture of 100 μl each of the two strains. Each plate was incubated statically for 18-20h, at 37 °C, with 5% oxygen in a plate reader (Tecan Spark M20), and OD_600_ was measured every 15 minutes. In order to explore the possibility that ill-defined components of THY may bias the differences seen, as well as to subject the cells to an environment more similar to the one found in the nasopharynx, chemically defined minimal media proposed by Aprianto *et al* [Aprianto *et al*., 2018] was tested, but the isolates failed to grow in this medium.

Curveball (ver. 0.2.5) was used to fit each growth curve to a bacterial growth model and calculate each strain-relative fitness by modelling the behaviour of each strain in co-culture. As curveball was designed to calculate fitness in *E. coli* growth experiments, the growth curves were censored after each strain reached the maximum OD_600_, to correct for the characteristic *S. pneumoniae* autolysis.

Association and invasion experiments on human respiratory tract epithelial cells (Detroit 562) were carried out as reported by Weight *et al*., 2019. Briefly, human pharyngeal carcinoma Detroit 562 epithelial cells (ATCC_CCL-138) were grown in 10% FCS in alpha MEM media (Gibco). Confluent Detroit 562 (typically day 8 post plating) were co-cultured with S*. pneumoniae* for 3 h in 1% FCS alpha MEM (MOI ∼1 cell:10 pneumococci). The medium was removed, and cells washed three times in Hanks Buffer Saline Solution (HBSS+/+, with calcium and magnesium, Gibco). Cells were incubated in 1% saponin for 10 min at 37 °C and lysed by repetitive pipetting. Dilutions of bacteria were plated on blood agar and colonies counted after 16 h. To quantify internalised bacteria, 100 µg/ml gentamicin was added for 1 h to the cells, which were washed another three times, before incubating with 1% Saponin and plating on blood agar plates. CFUs were counted after 16 h incubation at 37 °C, 5% CO_2_.

For testing the haemolytic activity, bacteria were suspended in phenol free RPMI (Invitrogen) and incubated with 2% red blood cells (EO labs) in a U-bottom 96 well plate for 30 minutes at 37°C / 5% CO_2_. Saponin was used as a positive control for cell lysis. The plate was centrifuged at 1500g for three minutes and supernatant was transferred to a new plate to read absorbance at 540nm.

Finally, Detroit 562 cells were infected with S. pneumoniae for 6 hours and supernatant was collected for analysis of IL-8 secretion. The protocol DuoSet® ELISA kit (R&D Systems) was followed according to manufacturers’ instructions.

## Data Availability

All required data are found in the Supplementary Material or archived online, as described in the Methods.

https://pubmlst.org/organisms/streptococcus-pneumoniae

## Acknowledgements

We thank the individuals who participated in this study and the local schools and authorities for their support. We are grateful to the study field teams (supported by Farouck Bonomali and Roseline Nyirenda) and the MLW laboratory team. We are grateful to the hospitality of the QECH ART Clinic, led by Ken Malisita. Our thanks also extend to the MLW laboratory management team (led by Brigitte Denis) and the MLW data management team (led by Clemens Masesa). R.S.H., N.F., A.G. and T.S. are supported by the National Institute for Health Research (NIHR) Global Health Research Unit on Mucosal Pathogens using UK aid from the UK Government. The views expressed in this publication are those of the author(s) and not necessarily those of the NIHR or the Department of Health and Social Care. We thank the High-Throughput Genomics Group at the Wellcome Trust Centre for Human Genetics (funded by Wellcome Trust grant reference 203141/Z/16/Z) for the generation of the Sequencing data.

## Author contributions

A.G., T.D.S., U.O., M.C.M., S.G., N.F., and R.S.H. designed the study and contributed to the development of the methodology. T.D.S., N.F., and R.S.H. oversaw the study, data collection, and data management. A.G., U.O., J.L. and S.G. developed genomic analysis. T. S., J. M. and C. B. oversaw the genome extraction and pneumococcal serotyping. A.G., U.O and T.D.S wrote the first draft of the paper. A.G., U.O., T.D.S., J.L., C.M.W., J.C., A.K., T.S.M., J.M., C.B., M.C.M., N.F., S.G., R.S.H. contributed to subsequent drafts and read and approved the final version of the report

## Ethics approval and consent to participate

The study protocol was approved by the College of Medicine Research and Ethics Committee, University of Malawi (P.02/15/1677) and the Liverpool School of Tropical Medicine Research Ethics Committee (14.056). Adult participants and parents/guardians of child participants provided written informed consent, children 8-10 years old provided informed assent. This included consent for publication.

## Supplementary figures and tables

**Figure S1 – frequency of STs, Serotypes and GPSCs in the three cohorts (vaccinated, unvaccinated children and adults).** In (a) and (c) the frequency of genotypes present in at least 10 strains is shown, in (b) the frequency of serotypes present in at least 2 strains is shown. The proportions shown in each plot are relative to the separate cohort.

**Figure S2 – Number of strains identified as a vaccine- or non-vaccine serotype, in common STs (a) or GPSC (b) in time.** STs or GPSCs present in more than 10 isolates are shown. Non-typeable serotype strain are excluded from this analysis.

**Figure S3 – (a) metabolic genotypes and (b) serotypes significantly different in distribution between survey 1-4 and survey 5-8. Statistical association between MT (c) or serotype (d) and penicillin resistance. (e) Penicillin resistance MIC in strains isolated during the early (survey 1-4) or late (survey 5-8) stages of the carriage surveys** . Panels a, b, c and d show volcano plots based on the p-value and the odds-ratio of fisher’s exact tests, calculating the significance of: each MT being differently distributed in survey 1-4 vs. 5-8 (a); each serotype being differently distributed in survey 1-4 vs. 5-8 (b); each strain in a MT showing a penicillin MIC higher than 0.06 ug/ml (c); each strain in a serotype showing a penicillin MIC higher than 0.06 ug/ml (d). Yellow and red lines show a significance of <0.01 (red) <0.05 (yellow). Dots are colored with the same color scheme.

**Figure S4 – Presence and absence of AMR genes in strains belonging to serotype 23B (a), 34 (b), 10A (c), 17F (d), 38 (e).** Each barplot shows the number of strains in which the specific AMR gene is present (+) or absent (-). Colors represent the metabolic genotype and are analogous to figure 3.

**Figure S5 - Core genome SNPs count for serotypes 38 (a), 17F (b), 34 (c), 10A (d), 23B (e).** Core genome SNPs were normalised against the lower SNP count in the core-genome alignment in each serotype [* F-test for equality of variances, p-value < 0.05]

**Figure S6 – Root-to-tip regression for strains isolated between 2015 and 2019 in Blantyre in the context of this study, for serotypes 38 (a) and 17F (b).** Dots on the tree are coloured from blue to red, according to their isolation date reflecting the dots on the linear regression plot. Nodes 85, 84 and 83 are highlighted in (a); Nodes 94 and node 95 are highlighted in (b)

**Figure S7 – Frequency of MTs, penicillin MIC and frequency of AMR genes in serotype 3 and 23F, during the 8 carriage surveys.** Each panel shows the frequency of isolation of each metabolic profile (connected points), penicillin MIC of each isolate (box and whiskers plot, with points representing each isolate), and presence of AMR genes in each metabolic type (regardless of isolation time - barplot). Panels correspond to serotypes 3 (top) and 23F (bottom). The box and whiskers plot in each panel also shows the number of isolates per survey at the top. Vertical blue lines separate the early-late isolates.

**Figure S8 - Core genome SNPs count for serotype 38 – MT 120 strains isolated in Malawi and in South Africa.** Core genome SNPs were normalised against the lower SNP count in the core-genome alignment in each serotype [* F-test for equality of 2 variances, p-value < 0.05]

**Figure S9 - Phylogenetic trees for post-vaccine carriage isolates of serotype (a) 10A, (b) 17F, (c) 38, (d) 23B, (e) 34**. Each tree is annotated with the metabolic genotypes (MT, colored strip) and with a binary heatmap showing gene presence (Green) or absence (Red) for the typical genes identified in the most common metabolic profile in the late stages of carriage surveys.

**Table S1 – GWAS analysis results**. For each serotype, the genes significatively enriched in the dominant MT are reported. Hypothetical proteins (as identified by automated Prokka annotation) are excluded.

**Table S2 –** Isolates sequenced in this study and associated metadata.

