## Supplementary Information for "The metabolic, virulence and antimicrobial resistance profiles of colonizing *Streptococcus pneumoniae* shift after pneumococcal vaccine introduction in urban Malawi"

**Figure S1 – frequency of STs, Serotypes and GPSCs in the three cohorts (vaccinated, unvaccinated children and adults).**

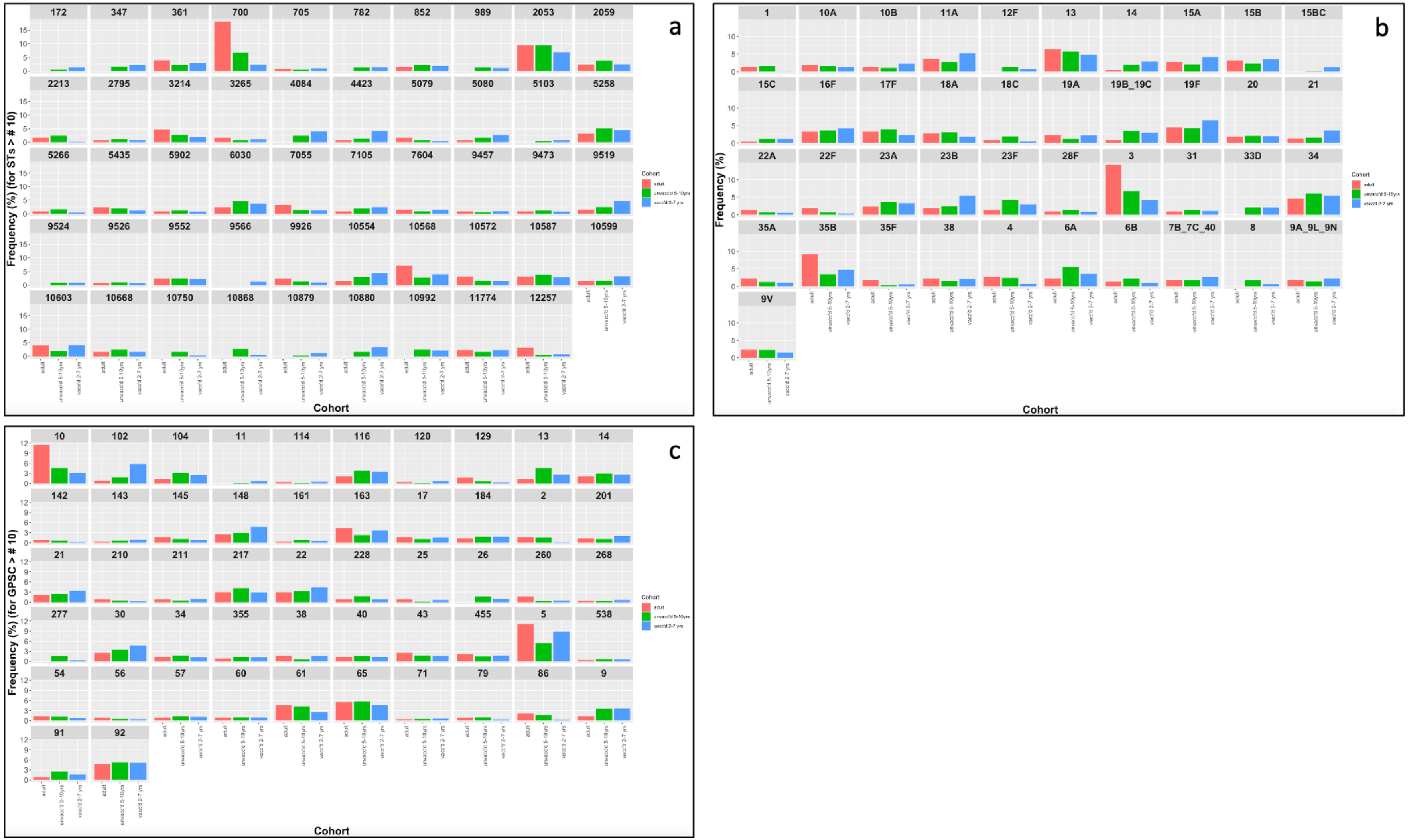

**Figure S2 – Number of strains identified as a vaccine- or non-vaccine serotype, in common STs (a) or GPSC (b) in time.**

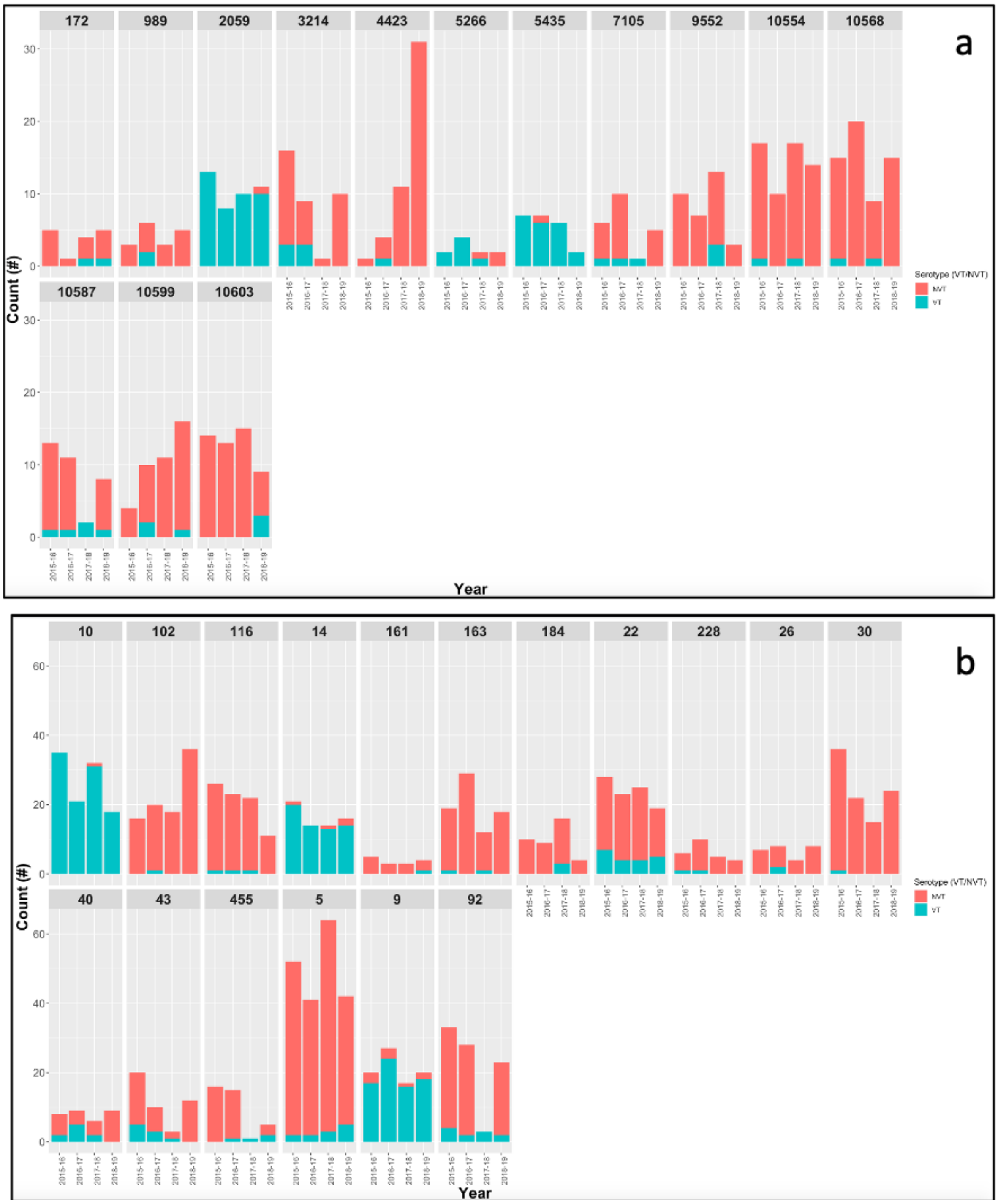

**Figure S3 – (a) metabolic genotypes and (b) serotypes significantly different in distribution between survey 1-4 and survey 5-6. Statistical association between MT (c) or serotype (d) and penicillin resistance. (e) Penicillin resistance MIC in strains isolated during the early (survey 1-4) or late (survey 5-6) stages of the carriage surveys.**

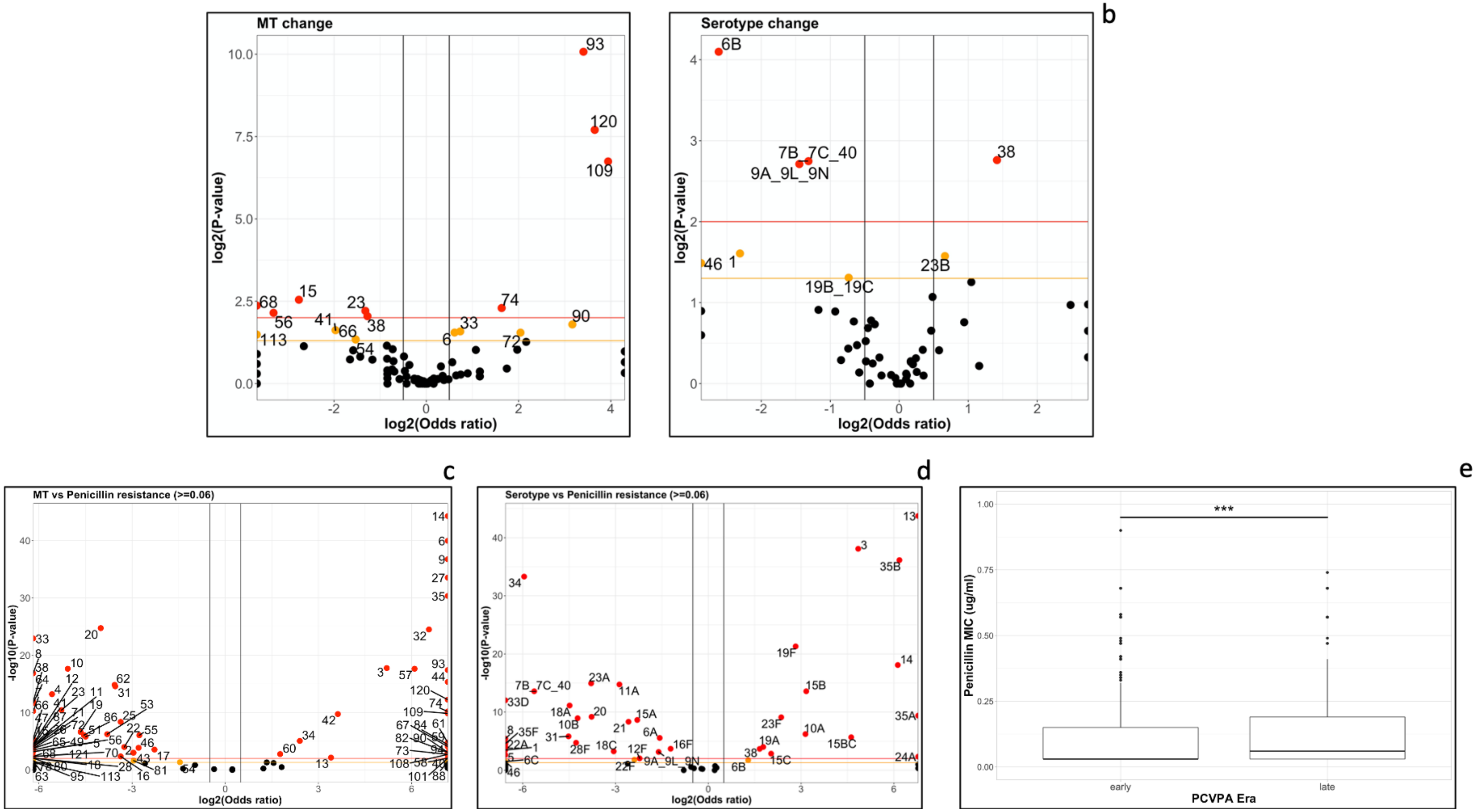

**Figure S4 – Presence and absence of AMR genes in strains belonging to serotype 23B (a), 34 (b), 10A (c), 17F (d), 38 (e).**

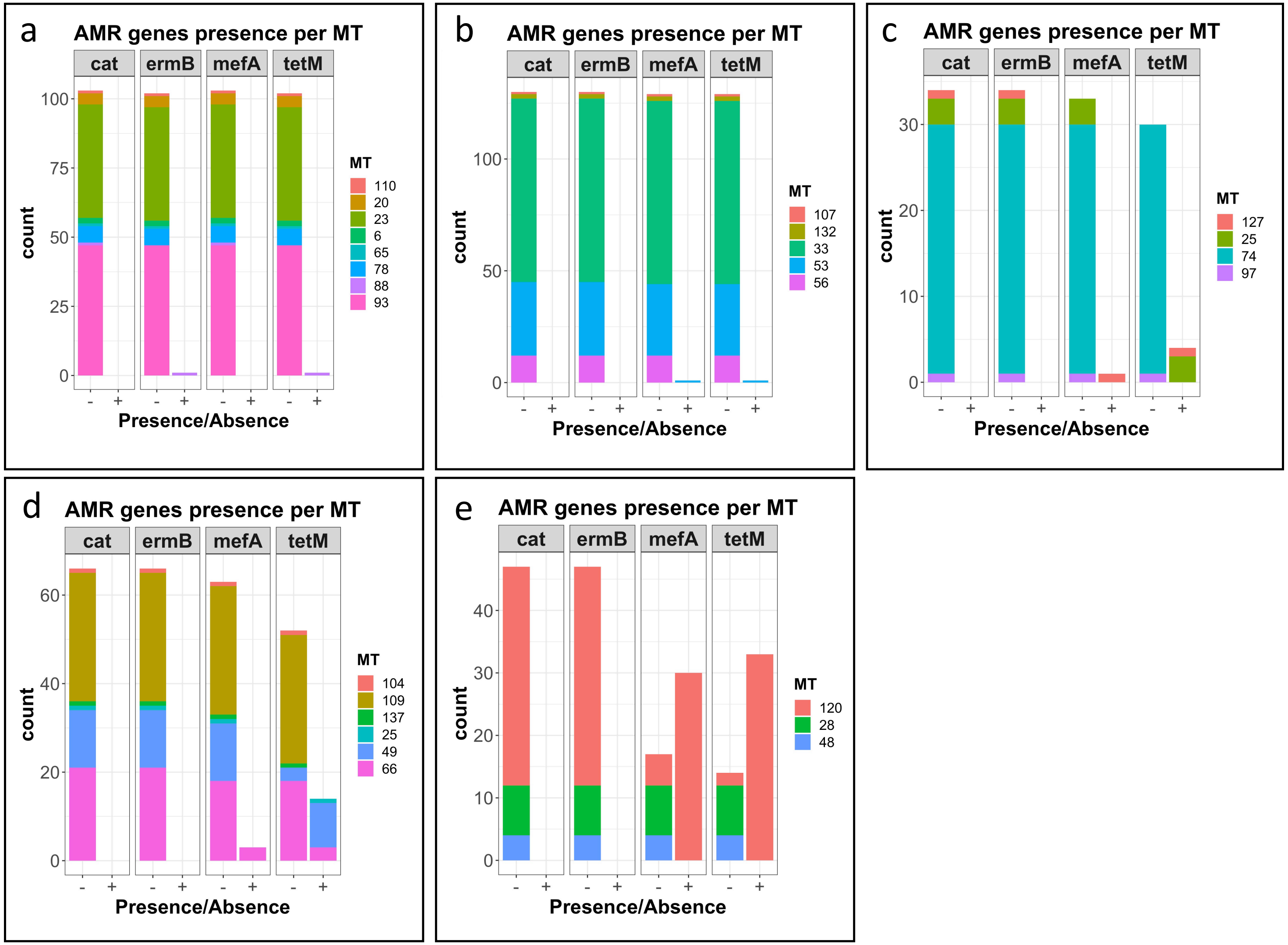

**Figure S5 – Frequency of MTs, penicillin MIC and frequency of AMR genes in serotype 3 and 23F, during the 8 carriage surveys.**

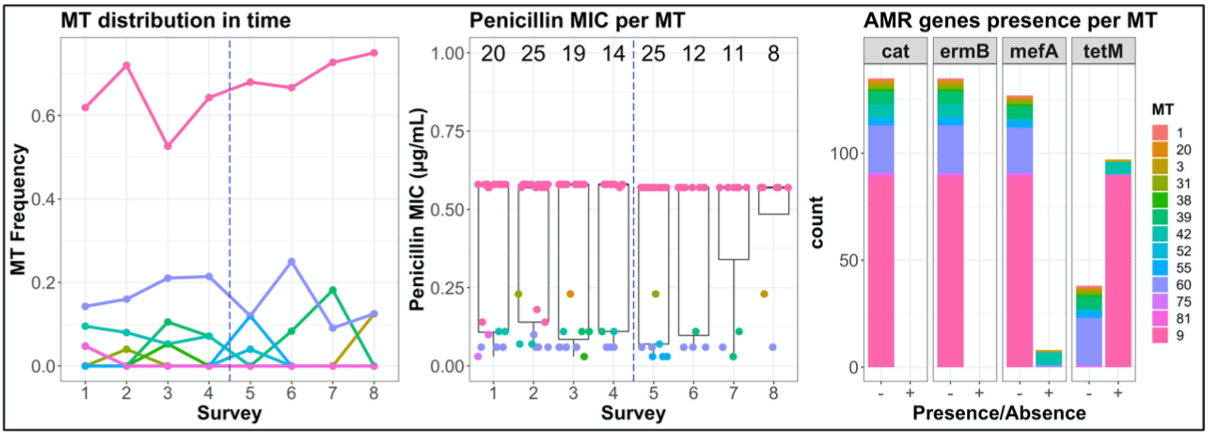


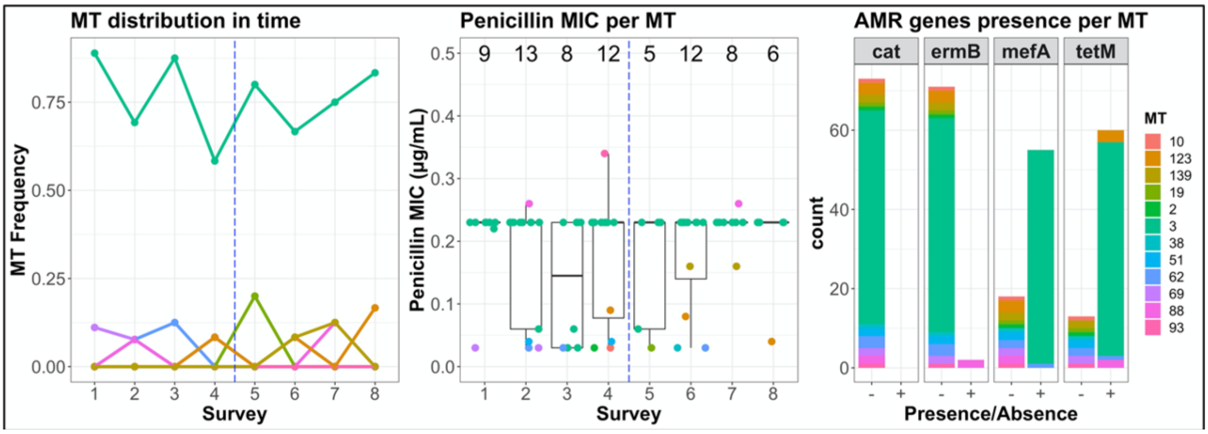

**Figure S6 - Core genome SNPs count for serotypes 38 (a), 17F (b), 34 (c), 10A (d), 23B (e).**

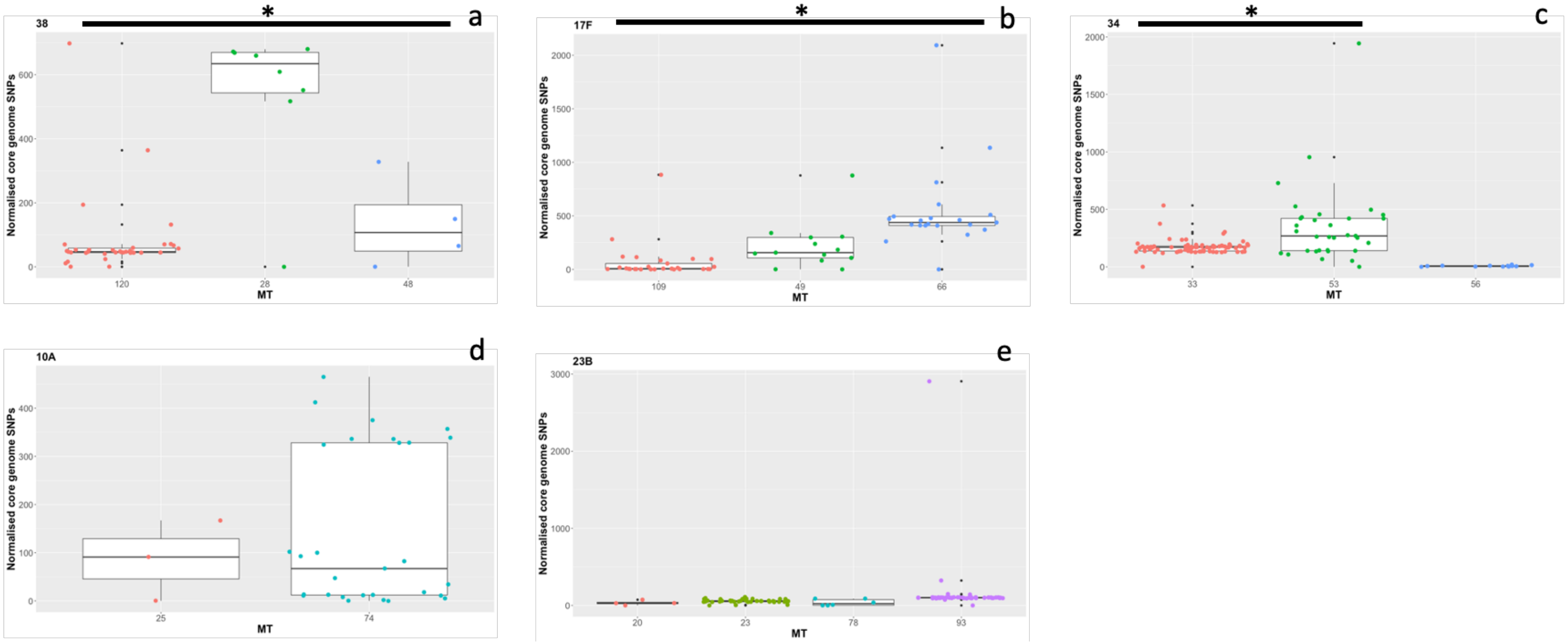

**Figure S7 – Root-to-tip regression for strains isolated between 2015 and 2019 in Blantyre in the context of this study, for serotypes 38 (a) and 17F (b).**

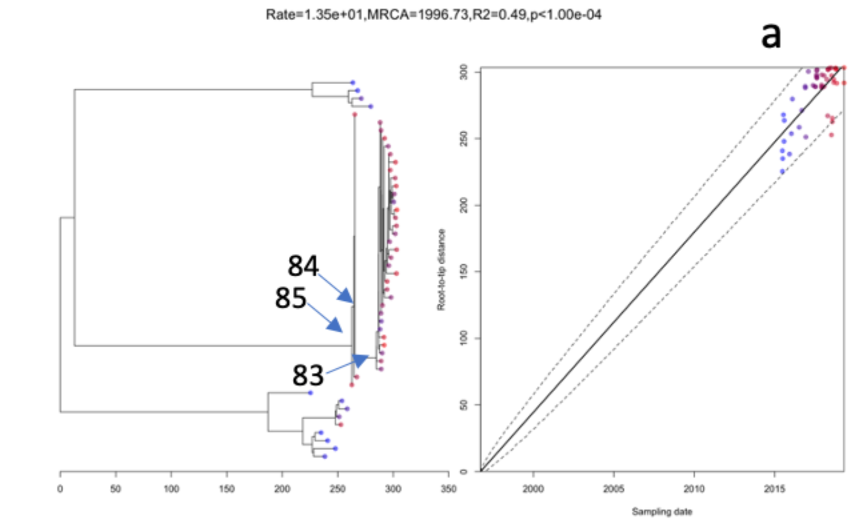


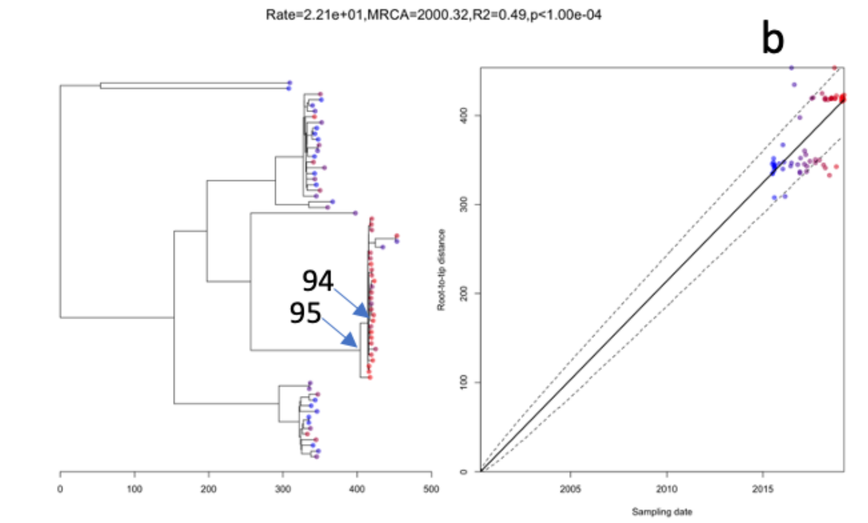

**Figure S8 - Phylogenetic trees for post-vaccine carriage isolates of serotype (a) 10A, (b) 17F, (c) 38, (d) 23B, (e) 34**

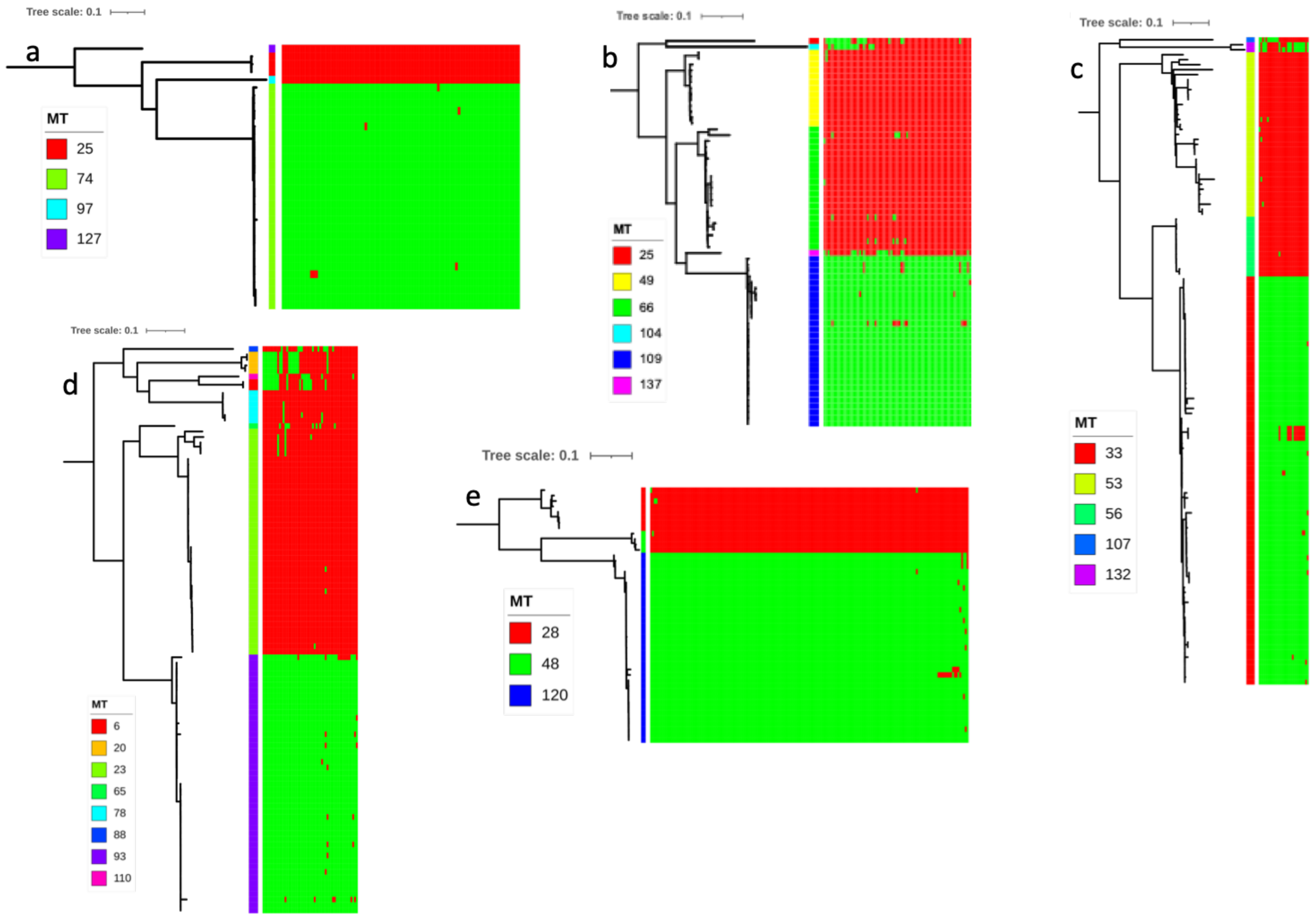

**Figure S9 - Core genome SNPs count for serotype 38 – MT 120 strains isolated in Malawi and in South Africa.**

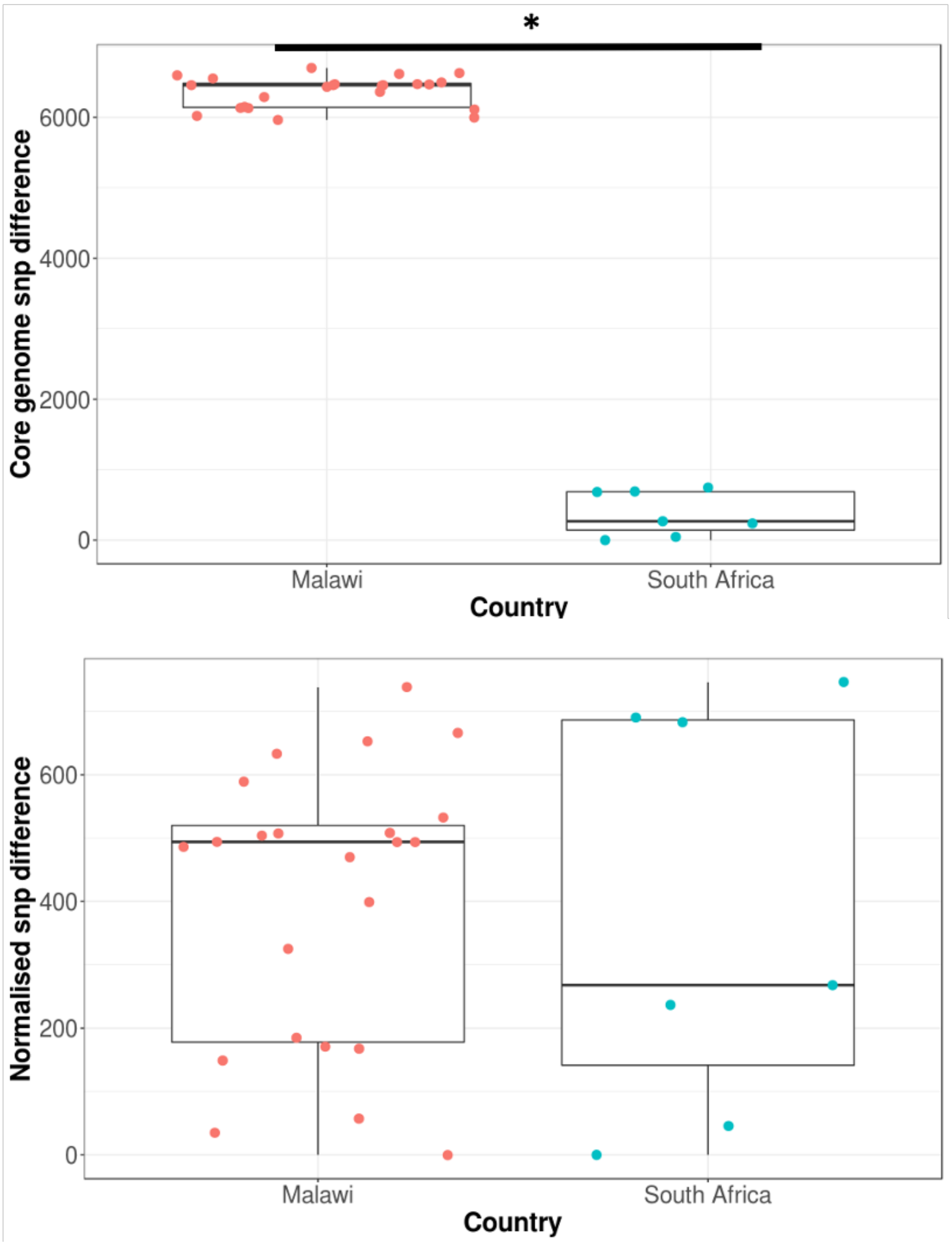
